# Transmission dynamics of Nipah virus in Bangladesh and India, 2001–2026: systematic review and inference on reproduction number, offspring dispersion, and serial interval

**DOI:** 10.64898/2026.07.16.26357631

**Authors:** Sol Kim, Vijayalaxmi V Mogasale, Juan F. Vesga, Hyolim Kang, Laura Skrip, Sung-mok Jung, Ausraful Islam, Akira Endo, W. John Edmunds, Kaja Abbas

## Abstract

**Background:** Nipah virus (NiV) is a priority zoonotic pathogen causing high-fatality outbreaks. Early NiV outbreaks in Malaysia and Singapore had limited transmission beyond spillover events. However, since 2001, NiV outbreaks with person-to-person transmission have occurred in Bangladesh and India, driven by the NiV-Bangladesh genotype and NiV-India genotype. Our study aims to estimate the reproduction number, offspring dispersion, and serial interval governing NiV transmission in Bangladesh and India during 2001–2026.

**Methods:** We conducted a systematic review of NiV outbreak investigations in Bangladesh and India, searching PubMed, Embase, Web of Science, and grey literature through 28 February 2026. Case-level offspring counts from 27 sources (323 cases across 67 outbreaks) were used as input to a hierarchical Bayesian negative binomial offspring distribution model. The serial interval was estimated by parametric distribution fitting to 137 transmission pairs. We conducted country-specific analysis and performed sensitivity analyses to evaluate the robustness of estimates.

**Results:** Pooling across 67 outbreaks, we estimated a median reproduction number of 0.43 (95% CrI: 0.26–0.75), an offspring dispersion parameter of 0.04 (0.03–0.06), and a serial interval of 13.3 days (95% CI: 12.8–13.8). Country-specific median reproduction numbers were 0.47 (0.22–0.98) for India and 0.35 (0.19–0.63) for Bangladesh, and dispersion parameters were 0.03 (0.02–0.06) and 0.05 (0.03–0.09), respectively, indicating marked overdispersion in both settings.

**Conclusion:** NiV transmission is self-limiting on average and highly overdispersed, suggesting that a disproportionate share of onward transmission arises from a small number of cases. This epidemiological profile supports targeted containment measures, including contact tracing and quarantine, for effective NiV outbreak control.

**Author Summary:** Nipah virus is a lethal infection that spreads from bats to people and, in some cases, from one person to another. Despite its severity as a high-consequence pathogen, there is no licensed vaccine or specific treatment. Since 2001, outbreaks have occurred almost every year in Bangladesh and India, which are the only places with person-to-person transmission. We gathered case reports from these outbreaks over twenty-five years and used a statistical model to measure how the virus spread between people. We found that, on average, each infected person passes the virus to fewer than one other person, so outbreaks tend to die out on their own without repeated introductions from animals. However, spread is heterogeneous: most people infect no one, while a small number infect many, occasionally producing large clusters. We estimated the typical time between one person falling ill and the next (serial interval) to be thirteen days. Together, these findings provide a better understanding of Nipah virus transmission and updated numbers needed to plan future prevention and control strategies for Nipah outbreaks.

## Introduction

Nipah virus (*Henipavirus nipahense*, NiV) is a zoonotic, negative-sense single-stranded RNA virus in the family *Paramyxoviridae*. It was first identified during the 1998–1999 Malaysia and Singapore outbreak, where transmission was primarily from infected pigs to pig farmers and abattoir workers (1,2). Since 2001, recurrent outbreaks in South Asia have been characterised by repeated zoonotic spillover from *Pteropus* fruit bats, followed by person-to-person transmission (3,4). In Bangladesh, the principal spillover route is consumption of raw or fermented date palm sap (‘tari’) contaminated by bat excretion, with additional contributions from contact with infected domestic animals such as pigs and cows (5–7). Three genotypes circulate: NiV-Malaysia (NiV-M), NiV-Bangladesh (NiV-B), and NiV-India (NiV-I) (8–10). NiV-I, closely related to NiV-B, has been associated with the recurring outbreaks in Kerala, India, since 2018 (8,10,11). Compared with NiV-M, infections caused by NiV-B and NiV-I are associated with higher case-fatality risks and more frequent respiratory manifestations (7,12,13). As of May 2024, at least 754 confirmed human cases had been documented across five countries in South and Southeast Asia, with overall case-fatality risks exceeding 70% in both Bangladesh and India, substantially higher than in the initial Malaysia and Singapore outbreaks (14,15).

NiV has been designated by the World Health Organization (WHO) and the Coalition for Epidemic Preparedness Innovations (CEPI) as a priority pathogen for research and development due to its epidemic potential (16–18). While no dedicated therapeutic or licensed vaccine is currently available for NiV, several vaccine candidates have shown promising Phase-1 immunogenicity data. Of note, ChAdOx1 NipahB is entering Phase-2 trials with up to 100,000 investigational doses planned for rapid outbreak deployment (19,20). Better epidemiological understanding of NiV will be critical for overall preparedness and for clinical trial planning for novel vaccine and therapeutic candidates.

Unlike the 1998–1999 Malaysia and Singapore outbreaks, which involved no person-to-person transmission and have not recurred, NiV outbreaks in Bangladesh and India have occurred almost every year since 2001 and are characterised by person-to-person transmission events. Bangladesh and India therefore represent the only settings in which the person-to-person transmission dynamics of NiV can be empirically studied, making them the focus for estimating epidemiological parameters relevant to pandemic risk assessment and vaccine deployment planning. Key parameters of interest characterise the offspring distribution, defined as the distribution of the number of secondary cases generated by a single infected individual. This is typically fit by applying a negative binomial distribution to available offspring distribution data (21).

The negative binomial offspring distribution is governed by the reproduction number (*R*) and the dispersion parameter (*k*). *R* is the mean of the offspring distribution and quantifies average transmissibility. It defines the epidemic threshold: transmission is self-sustaining when *R* ≥ *1* and subcritical, tending to extinction, when *R* < *1*. The dispersion parameter (*k*) governs how unevenly transmission is distributed across cases—when *k* is large, most cases transmit similarly to the average; when *k* is small, the majority of cases generate no or very few secondary infections while a minority are responsible for most onward transmission. Together with the serial interval, the interval between symptom onset in an infector and the person they infect (infectee), these parameters characterise the transmission dynamics that inform decisions on the prevention and control of Nipah outbreaks and plan for implementation strategies for new NiV vaccines in the development pipeline.

To date, published estimates of both parameters derive almost exclusively from Bangladesh and are lacking for India (6,15,22–24). To address this evidence gap, we systematically reviewed publicly available literature on NiV outbreaks in Bangladesh and India, and estimated the reproduction number, offspring dispersion, and serial interval of NiV transmission during 2001–2026.

## Methods

### Search strategy and study selection

We searched PubMed, Embase, and Web of Science through February 28, 2026, using terms related to Nipah virus and epidemiology, with no date or language restrictions (Supplementary Table S2). WHO Disease Outbreak News and the icddr,b Health and Science Bulletin were also searched as additional sources (25,26). The review is registered with PROSPERO (CRD42025641668).

We included studies that reported observational human NiV outbreaks. We excluded studies restricted to animals, systematic reviews and other review articles, and modelling studies (Supplementary Table S3). Title and abstract screening was performed by one reviewer (SK); full-text review was conducted independently by two reviewers (SK, VVM), with disagreements resolved by discussion and consensus.

### Data extraction

Data were extracted by one reviewer (SK) and independently verified by a second reviewer (VVM), using a standardised data extraction spreadsheet, on study year, setting, infector-infectee pairs, dates of symptom onset per case, and other variables (Supplementary Table S4). When multiple studies reported the same outbreak, we prioritised the source with the more complete case ascertainment and detailed infector-infectee data (Supplementary Text S1). For this analysis, an outbreak was defined as one or more confirmed or probable NiV case(s)—classified according to the case definitions applied in the original source study—that were geographically and temporally clustered.

### Quality assessment

Risk of bias was assessed by two independent reviewers (SK, VVM) using JBI critical appraisal tools for case-control studies, case series, and case reports (Supplementary Table S6) (27). Discrepancies were resolved by discussion.

### Transmission pair reconstruction

Transmission pairs were reconstructed from the included outbreak investigation reports at the case level, recording infector–infectee links, symptom onset dates, and secondary case counts. Each human case was assigned an offspring count equal to the number of person-to-person transmission events in which they subsequently appeared as the infector. Cases generating no further transmission were assigned a null offspring count. Non-human or environmental sources (e.g., bats, date palm sap) appearing only as primary infectors were not used to construct the offspring distribution (Supplementary Texts S2; Text S3).

### Offspring distribution model

To better account for variation in transmission context across settings, time periods, and countries, we fitted a hierarchical negative binomial model in which each outbreak *j* has its own reproduction number (*R_j_*) drawn from a lognormal distribution, while a single dispersion parameter *k* is shared across all cases and outbreaks. This structure allows the reproduction number to vary across outbreaks while partially pooling information across them, and separates two distinct sources of variation in the data: individual-case heterogeneity in transmission (*k*) and between-outbreak heterogeneity in reproduction number (*σ_R_*, the log-scale standard deviation of the lognormal distribution). The model is specified as follows:

For case *i* in outbreak *j*, the number of secondary cases *X_ij_* follows:

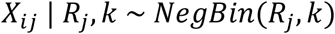

where *i* = *1*, . . . , *n_j_* index cases for *n_j_* cumulative cases in outbreak *j*. On the log scale:

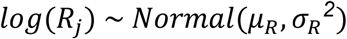

where *μ_R_* is the mean of log(*R_j_*) across outbreaks. From the fitted lognormal distribution of outbreak-specific reproduction numbers, two summary measures were derived: the median, *R* (median) = exp(*μ_R_* ), representing the reproduction number of a typical outbreak and used as the primary estimate throughout; and the arithmetic mean, *R* (mean) = exp(*μ_R_* + *σ_R_^2^*/*2*), the average reproduction number across outbreaks, reported for downstream modelling applications. Both quantities were computed for each posterior draw and summarised as posterior medians with 95% credible intervals.

Three prior specifications (P1: weakly informative reference; P2: epidemiologically informed, permissive between-outbreak heterogeneity; and P3: epidemiologically informed, conservative between-outbreak heterogeneity) were evaluated. Parameters were estimated in a Bayesian framework using the No-U-Turn Sampler (NUTS) in Stan via the *cmdstanr* package in R (version 4.5.2), using a non-centred parameterisation (28–30). Full prior specifications and sampler settings are provided in Supplementary Text S4, and posterior predictive check definitions in Supplementary Text S6.

Country-specific analyses fitted the same hierarchical negative binomial model independently to Bangladesh and India subsets under P2 (epidemiologically informed, permissive between-outbreak heterogeneity) prior specification. Between-country differences were assessed by contrasting the independent country posteriors on the log scale, summarised as the posterior probability of difference and the median India-to-Bangladesh ratio (95% credible interval). Sensitivity analyses examined: (1) restriction to outbreaks with person-to-person transmission (SA-O-P2P); (2) exclusion of the two manually inferred index cases (SA-O-MI) (Supplementary Text S3); (3) Bangladesh surveillance-era stratification by pre- and post-2007 periods (2001–2006 vs. 2007 onwards), corresponding to the introduction of hospital-based sentinel surveillance (SA-O-B1, SA-O-B2); (4) For Bangladesh, we set 137 cases to represent uncaptured single-case spillovers, derived from the difference between the 211 cases in the current dataset and the 348 cases reported by the Institute of Epidemiology, Disease Control and Research (IEDCR) (SA-O-B3) (31); and (5) we reparameterised the variance-to-mean ratio to hold constant across outbreaks, with *ψ* = *R*/*k* shared in place of *k* (SA-O-CV). Methodological details of the sensitivity analyses are provided in the Appendix (Supplementary Text S5).

### Serial interval estimation

The serial interval was defined as the number of days between symptom onset in the infector and symptom onset in the infectee. Pairs were included if both onset dates were available; no negative serial intervals were present in the data. Gamma, lognormal, and Weibull distributions were fitted by maximum likelihood using the *fitdistrplus* package in R, with distribution selection based on Akaike Information Criteria (AIC) (32,33). Uncertainty in the fitted mean and standard deviation was quantified by non-parametric bootstrap with 1,000 resamples, refitting the selected distribution within each replicate. Country-specific fitting was conducted independently within each subset with independent distribution selection. Two sensitivity analyses (referred to as SA-S1 and SA-S2) were conducted. The first restricted the analysis to confirmed transmission pairs by excluding infectees with multiple candidate infectors (SA-S1). The second involved a separate analysis of Bangladesh transmission pairs from pre- and post-2007 periods to assess the sensitivity of the estimate to surveillance changes (SA-S2).

## Results

### Study selection and quality assessment

A total of 2,686 unique articles were identified through the review of published literature, and 26 additional data sources were identified through repositories, grey literature sources, and review of reference lists (Figure 1). 34 references met the inclusion criteria after full-text review (25,26,34). This included one WHO SEARO Nipah Webinar report (2026) which served as a supporting data source for the 2025– 2026 West Bengal outbreak (35). During data extraction, seven studies were excluded because they reported the same outbreaks with less complete case ascertainment than the selected primary sources (Supplementary Text S1). Ultimately, 27 sources were used for data extraction (Supplementary Table S5).

**Figure 1.**
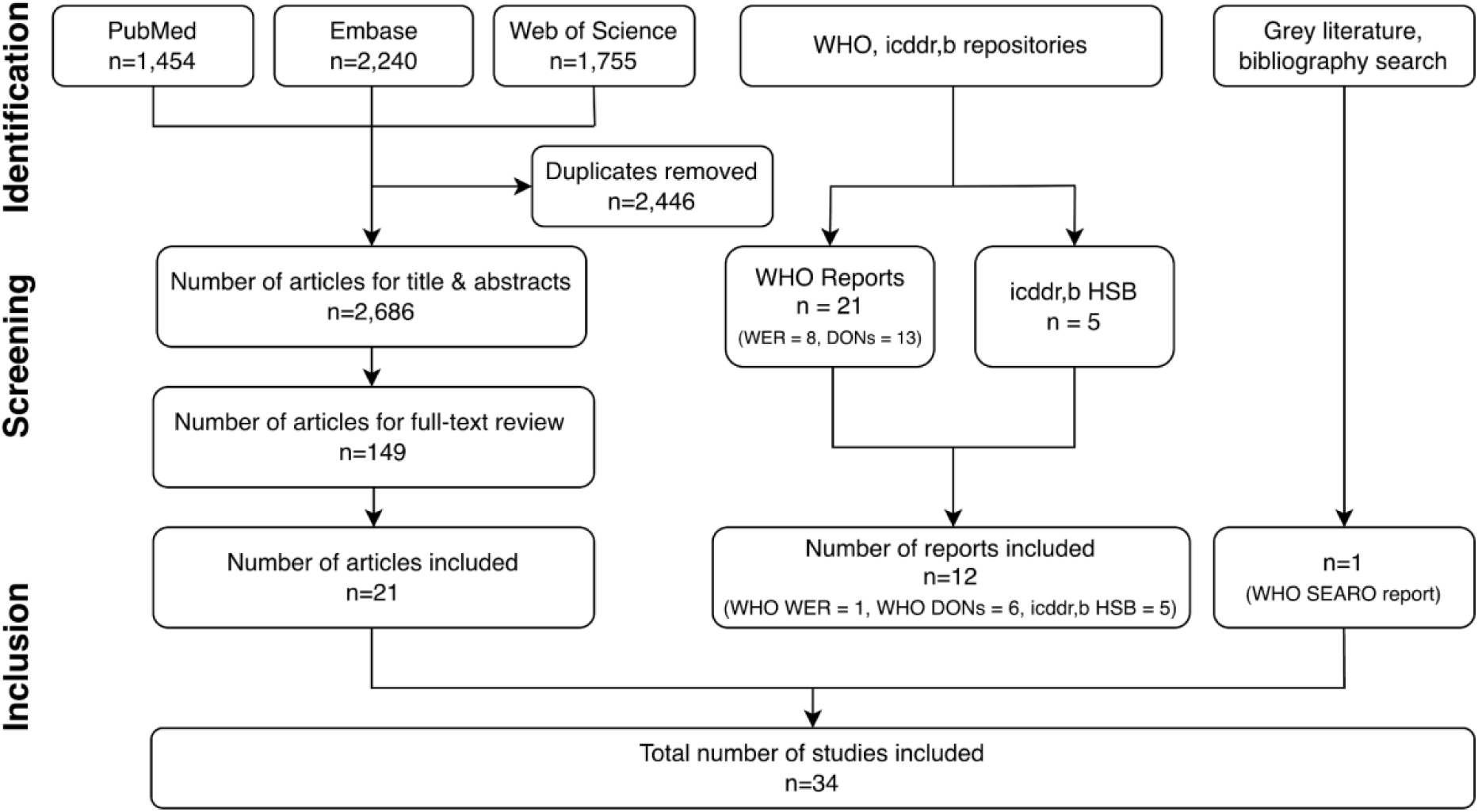
Systematic review of Nipah virus outbreaks in Bangladesh and India. PRISMA (Preferred Reporting Items for Systematic reviews and Meta-Analyses) flow diagram showing the number of records at each stage of identification, screening, and inclusion.

The included sources comprised nine case-control studies (5,36–43), 10 case series (11,44–52), and two case reports (53,54). Across study types, included references were generally considered of moderate or high quality, with one exception (11). The most consistently underreported items across study types were consecutive and complete inclusion of participants and identification and management of confounding (Supplementary Table S6).

### Outbreak summary

The 34 articles documented 67 discrete outbreaks spanning 2001–2026 (Table 1): 54 in Bangladesh (211 cases including 166 deaths) and 13 in India (112 cases including 80 deaths) (Figure 2). Outbreaks were classified into three types: (1) clusters with at least one documented person-to-person transmission pair (n = 21 outbreaks with 245 cases); (2) single-case spillover events with no onward transmission (n = 40 outbreaks); and (3) multi-case spillover clusters with no person-to-person transmission involved, in which cases occurred in the same area within a defined period, suggesting a possible shared zoonotic source such as a common contaminated date palm sap (n = 6 outbreaks with 38 cases) (Supplementary Figure S1). Transmission chain depth, defined as the number of person-to-person transmission events following the index case (Generation 0), was characterised for the 21 outbreaks with documented person-to-person transmission (16 outbreaks, 141 cases, in Bangladesh; five outbreaks, 104 cases, in India) (Supplementary Table S7). Most outbreaks (17/21, 81%) were limited to a single transmission generation beyond the index case (depth = 1); four (19%) extended further, with a maximum depth of four (five generations; 2004 Faridpur). In Bangladesh, 14 of 16 outbreaks (87.5%) reached depth 1, and the remaining two outbreaks reached depths of two (2001 Meherpur) and four (2004 Faridpur). In India, three of five outbreaks (60%) reached depth 1, and the remaining two outbreaks reached a depth of two (2001 Siliguri and 2018 Kerala).

**Figure 2.**
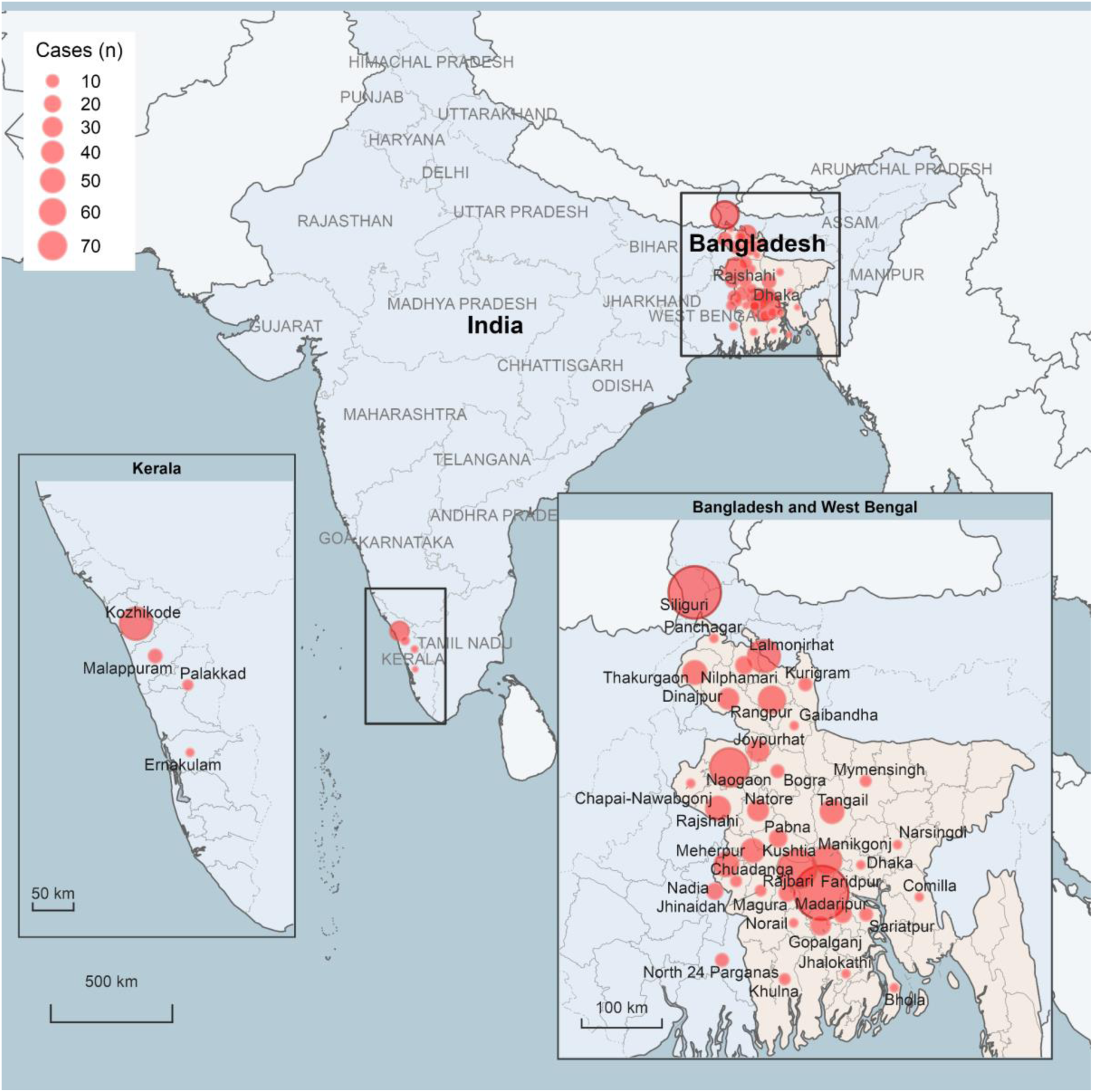
Geographic distribution of Nipah virus outbreaks in Bangladesh and India. Point size reflects the total number of cases per district from 2001 to 2026. Bangladesh totals during 2001-2026 are from the Institute of Epidemiology, Disease Control and Research (IEDCR) district-year counts (348 cases in 35 districts; https://iedcr.gov.bd/). India totals are retrieved by the present study (111 cases in 7 districts). Cases with unknown location are not shown on the map [n = 1, India (2018)]. Locations are district centroids. Natural Earth 1:10m Cultural Vectors (public domain; naturalearthdata.com). District boundaries are geoBoundaries gbOpen ADM2, CC BY 4.0 (geoboundaries.org).

**Table 1.**
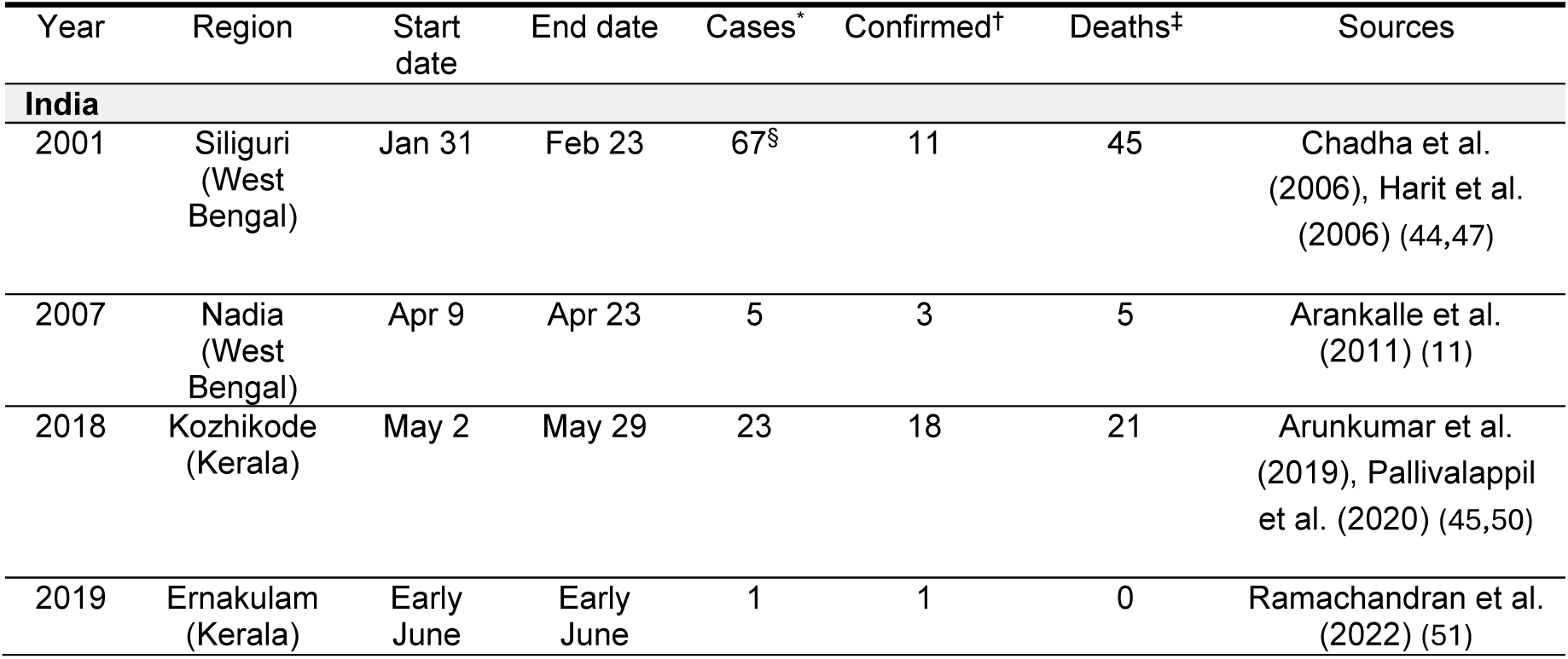
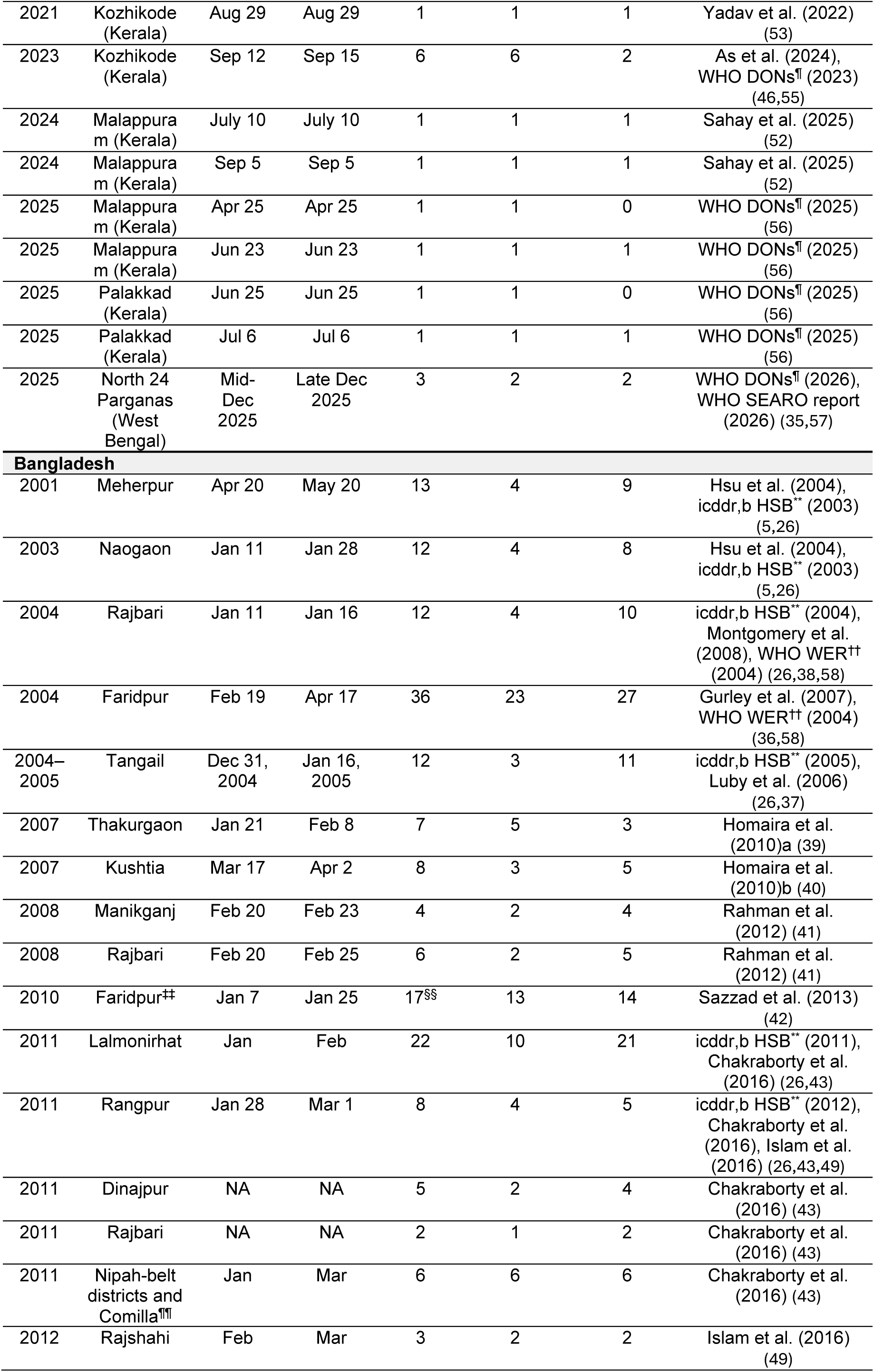
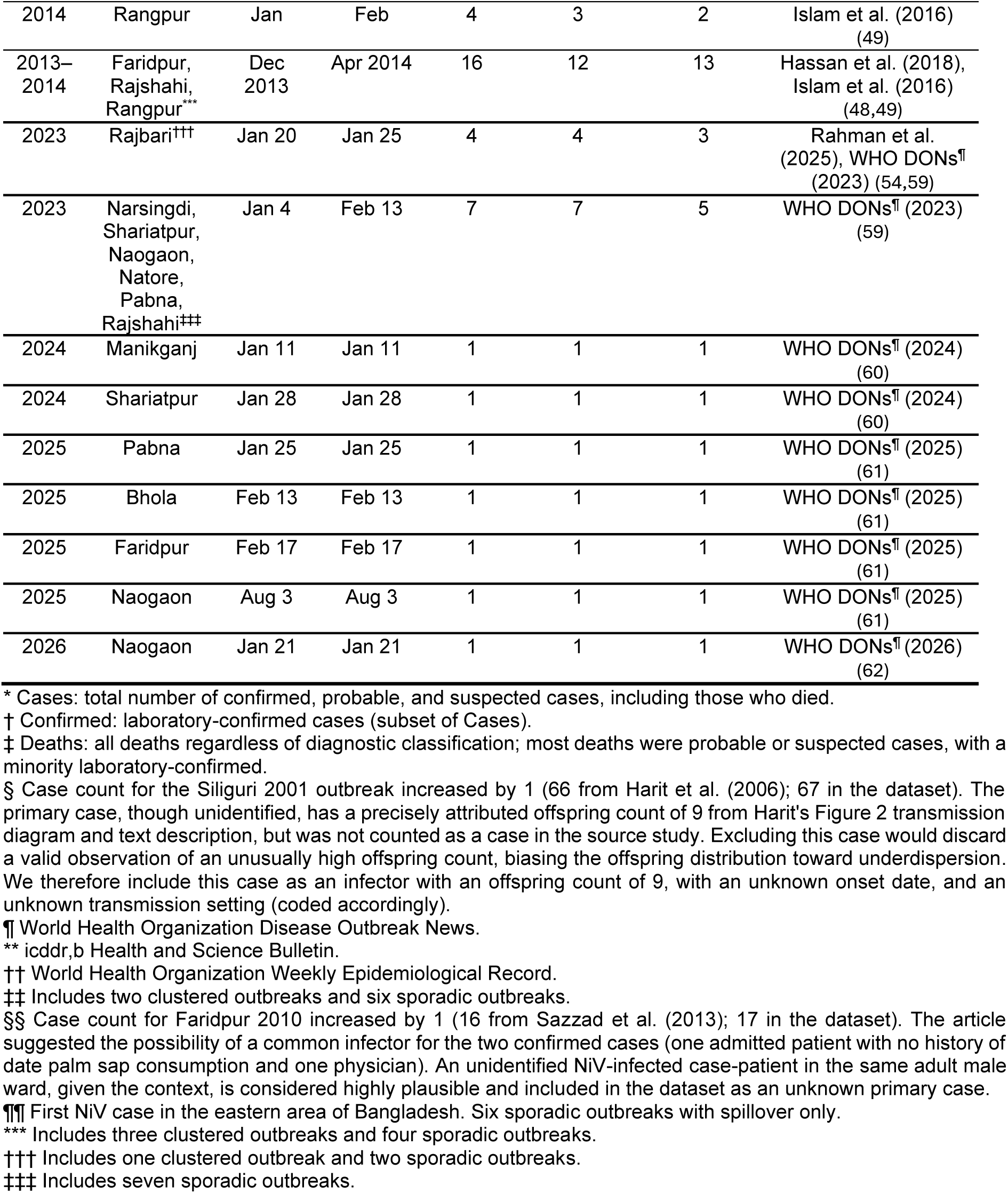
Nipah virus outbreaks in India and Bangladesh.

We also examined the setting in which person-to-person infection occurred. Of the 245 cases in these 21 person-to-person outbreaks, 157 (64%) were secondary cases (generation ≥ 1), and 88 (36%) were generation 0 index or spillover cases. Among the 157 person-to-person secondary cases, 49% were infected in hospital, 21% in households, 19% in an unknown setting, and 11% in the community. This pattern differed sharply by country: 93% of Indian person-to-person infectees were hospital-acquired (71/76), compared with 7% in Bangladesh (6/81). Most infections occurred at generation 1 (90/157). Hospital transmission contributed many cases at generations 1–2 but did not produce the longest chains. The only outbreak with depth 4 (2004 Faridpur) was community- and unknown-dominated. Household infections were almost always a single step from the index (94% at generation 1) and rarely led to further transmission.

For Bangladesh, the 211 cases ascertained for transmission pair reconstruction in this analysis represent 61% (211/348) of the 348 cases reported by surveillance data for the full 2001–2026 (February) period (Figure 3) (7,14). Data on the remaining cases could not be ascertained since discrete outbreak investigation reports during 2015– 2022 were not identified from publicly accessible publications (6,7). While the number of cases reported through the Bangladesh surveillance system is publicly available, that source does not provide outbreak-level information required for further analysis.

**Figure 3.**
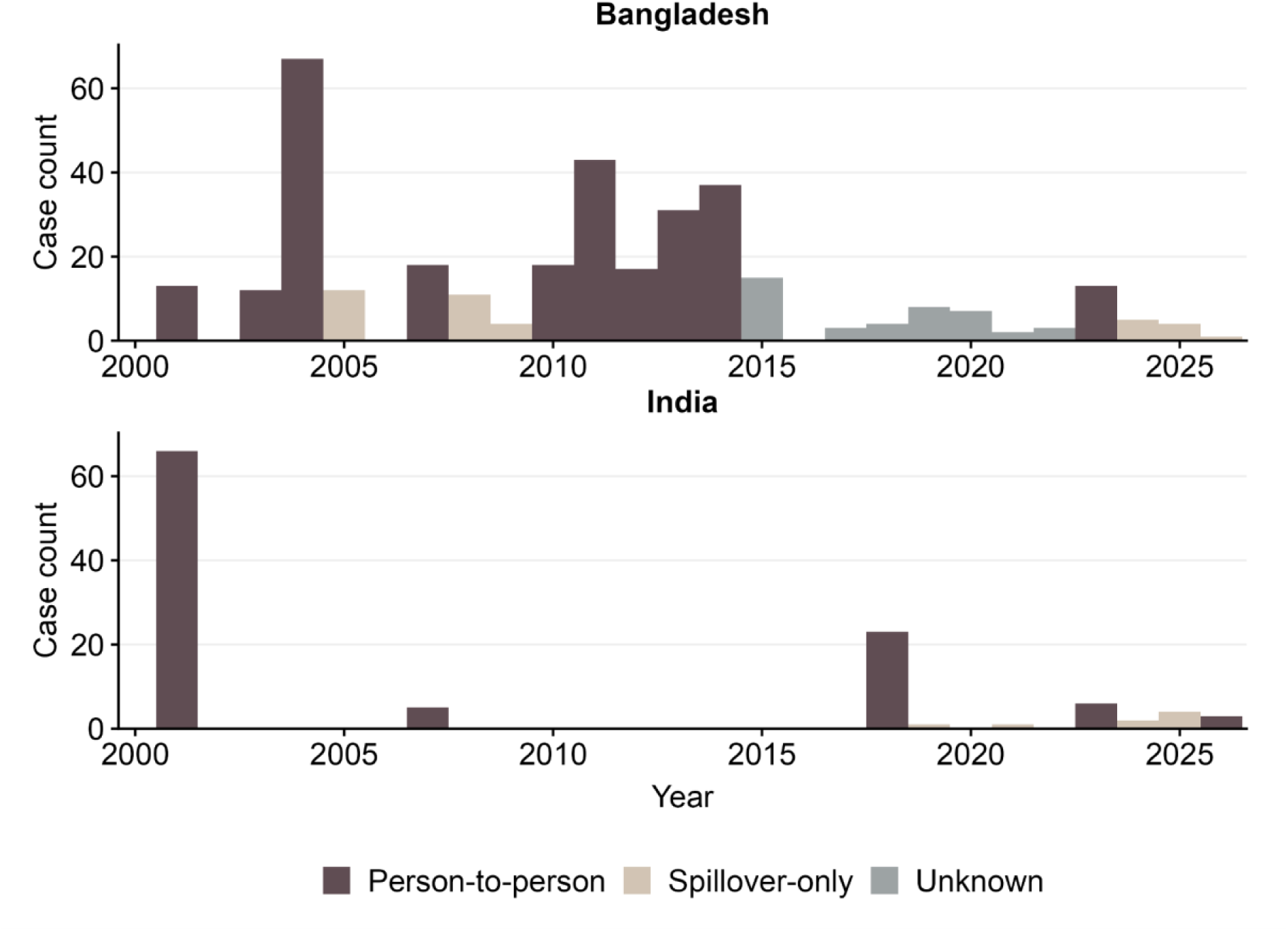
Nipah virus case counts, Bangladesh and India, 2001–2026. Confirmed and probable Nipah virus cases per year are shown. Colour indicates whether person-to-person transmission was documented in at least one outbreak in that year (dark taupe), all outbreaks were single-spillover cases (pale taupe), or transmission classification could not be determined from available sources (grey; 2015–2022, Bangladesh; data source: Institute of Epidemiology, Disease Control and Research (IEDCR), https://iedcr.gov.bd/).

### Reproduction number and offspring dispersion

Across 323 cases from 67 outbreaks, the hierarchical negative binomial model under prior set P2 (epidemiologically informed, permissive between-outbreak heterogeneity) yielded median *R* = 0.43 (95% CrI: 0.26–0.75) (Table 2). The mean *R*, reported for downstream modelling applications requiring an average over the outbreak distribution, was 0.47 (0.28–0.90). The offspring dispersion parameter *k* = 0.04 (0.03– 0.06) indicates high individual-level heterogeneity in transmission (Table 2). Under priors P1–P3, estimates of median *R* and *k* were largely data-driven, whereas the between-outbreak heterogeneity parameter *σ_R_* was weakly identified, with a credible interval that could not distinguish negligible from substantial between-outbreak heterogeneity (Table 2). Details on prior sensitivity and model validation results are provided in the Appendix (Supplementary Text S4; Text S6; Table S8; Figure S2–S5).

**Table 2.** Transmission parameter estimates for reproduction number, offspring dispersion parameter, and serial interval.

| Stratum | Outbreaks | Cases | Reproduction number<br>$R$ (median)<br>[95% CrI] | Reproduction number<br>$R$ (mean)<br>[95% CrI] | Between-outbreak heterogeneity<br>$\sigma_R$<br>[95% CrI] | Offspring dispersion parameter<br>$k$<br>[95% CrI] | Serial Interval mean, days<br>[95% CI]<br>(n pairs) |
| --- | --- | --- | --- | --- | --- | --- | --- |
| Pooled | 67 | 323 | 0.43 [0.26–0.75] | 0.47 [0.28–0.90] | 0.28 [0.01–1.00] | 0.04 [0.03–0.06] | 13.3<br>[12.8–13.8]<br>(n = 137) |
| Bangladesh | 54 | 211* | 0.35 [0.19–0.63] | 0.40 [0.23–0.87] | 0.39 [0.02–1.24] | 0.05 [0.03–0.09] | 12.8<br>[12.0–13.5]<br>(n = 70) |
| India | 13 | 112** | 0.47 [0.22–0.98] | 0.55 [0.27–1.75] | 0.42 [0.02–1.59] | 0.03 [0.02–0.06] | 13.9<br>[13.2–14.5]<br>(n = 67) |

#### Country-specific analyses

Fitting each country separately, the median *R* was 0.47 (0.22–0.98) in India and 0.35 (0.19–0.63) in Bangladesh, and the dispersion parameter *k* was 0.03 (0.02–0.06) and 0.05 (0.03–0.09), respectively (Table 2; Figure 4). Both countries showed high overdispersion (*k* ≪ *1*). The posterior probability that *R* was higher in India than Bangladesh was 0.73 (median ratio 1.33 [0.51–3.42]), and the probability of lower *k* in India was 0.85 (median ratio 0.65 [0.26–1.48]). Neither contrast was conclusive. *σ_R_* was prior-sensitive in both countries. India’s median *R* was sensitive to prior specification, ranging from 0.47 (0.22–0.98) under P2 to 0.84 (0.30–3.04) under P1, whereas P2 and P3 gave concordant estimates (0.47 and 0.48), suggesting the India data are insufficient to constrain *R* without informative prior structure. Bangladesh’s *R* was more stable across priors (0.35–0.42). Both countries’ *k* estimates were robust across all prior specifications. More details on country-specific estimates and posterior predictive checks are provided in the Appendix (Supplementary Text S6;_Table S9; Figure S2; and Figure S6).

**Figure 4.**
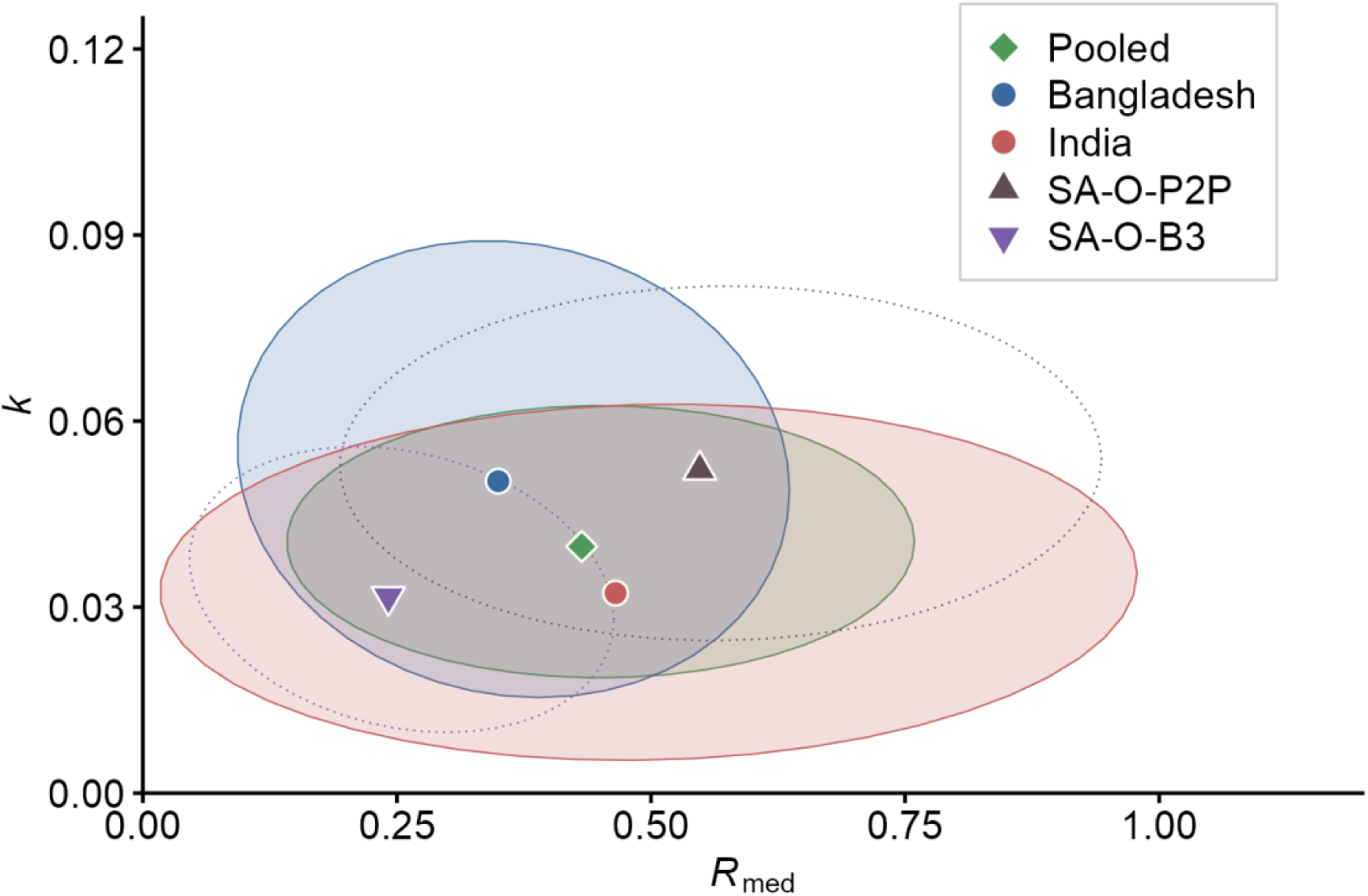
Reproduction number and offspring dispersion. Posterior medians of the reproduction number (*R_med_*) and the offspring dispersion parameter (*k*) from independent hierarchical negative binomial fits under prior set P2 (epidemiologically informed, permissive between-outbreak heterogeneity). Diamond: pooled analysis (67 outbreaks, 323 cases). Circles: country-specific analyses (Bangladesh, 54 outbreaks, 211 cases; India, 13 outbreaks, 112 cases). Triangle: sensitivity analysis restricted to person-to-person outbreaks (21 outbreaks, 245 cases; SA-O-P2P). Inverted triangle: sensitivity analysis with augmented Bangladesh dataset (Bangladesh, 191 outbreaks, 348 cases; SA-O-B3), a conservative lower bound on *R_med_*. Ellipses denote 95% bivariate normal credible regions of the joint posterior.

#### Sensitivity analyses

Across all structural and parameterisation sensitivity analyses, *R* (median) remained below 1 and *k* below 0.2 (Figure 4, Table 3). Restriction to outbreaks with person-to-person transmission (SA-O-P2P; 21 outbreaks; 245 cases) yielded *R* (median) = 0.55 (0.33–0.93) and *k* = 0.05 (0.03–0.08). This estimate characterises person-to-person transmissibility, providing an upper bound for transmission potential while remaining subcritical. Excluding the two manually inferred index cases from 2001 Siliguri and 2010 Faridpur outbreaks (67 outbreaks, 321 cases; SA-O-MI) yielded *R* (median) = 0.41 (0.24–0.71) and *k* = 0.04 (0.02–0.06), closely mirroring primary model estimates (*R* (median) = 0.43 [0.26–0.75]; *k* = 0.04 [0.03–0.06]) and suggesting that transmission tree reconstruction choices do not drive the overdispersion estimate. Stratification of the Bangladesh dataset by surveillance era yielded a median *R* of 0.39 (0.17–0.87) in the pre-surveillance period (SA-O-B1; 5 outbreaks, 85 cases) and 0.28 (0.15–0.57) under prospective surveillance from 2007 onward (SA-O-B2; 49 outbreaks, 126 cases). Augmenting the Bangladesh analysis with 137 additional national surveillance cases (IEDCR) (31), assumed to be non-transmitting spillovers (SA-O-B3; 348 cases covering from 2001 to 2026) established a conservative lower bound of *R* (median) = 0.24 (0.13–0.45) and *k* = 0.03 (0.02–0.05), compared with *R* (median) = 0.35 (0.19–0.63) and *k* = 0.05 (0.03–0.09) in the Bangladesh country analysis (Table 2). Reparameterising the offspring variance so that the variance-to-mean ratio was held constant across outbreaks (SA-O-CV) yielded *R* (median) = 0.40 (0.25–0.66) and an implied *k* at the median *R* of 0.04 (0.02–0.06). The fitted *ψ* = 10.22 (5.90–19.08) closely matched the ratio implied by the primary model posterior (*ψ derived* = 10.87 [5.72–22.00]), consistent with the conclusion that subcritical transmission and overdispersion were robust to the dispersion specification (Table S10–S11; Supplementary Text S6).

**Table 3.**
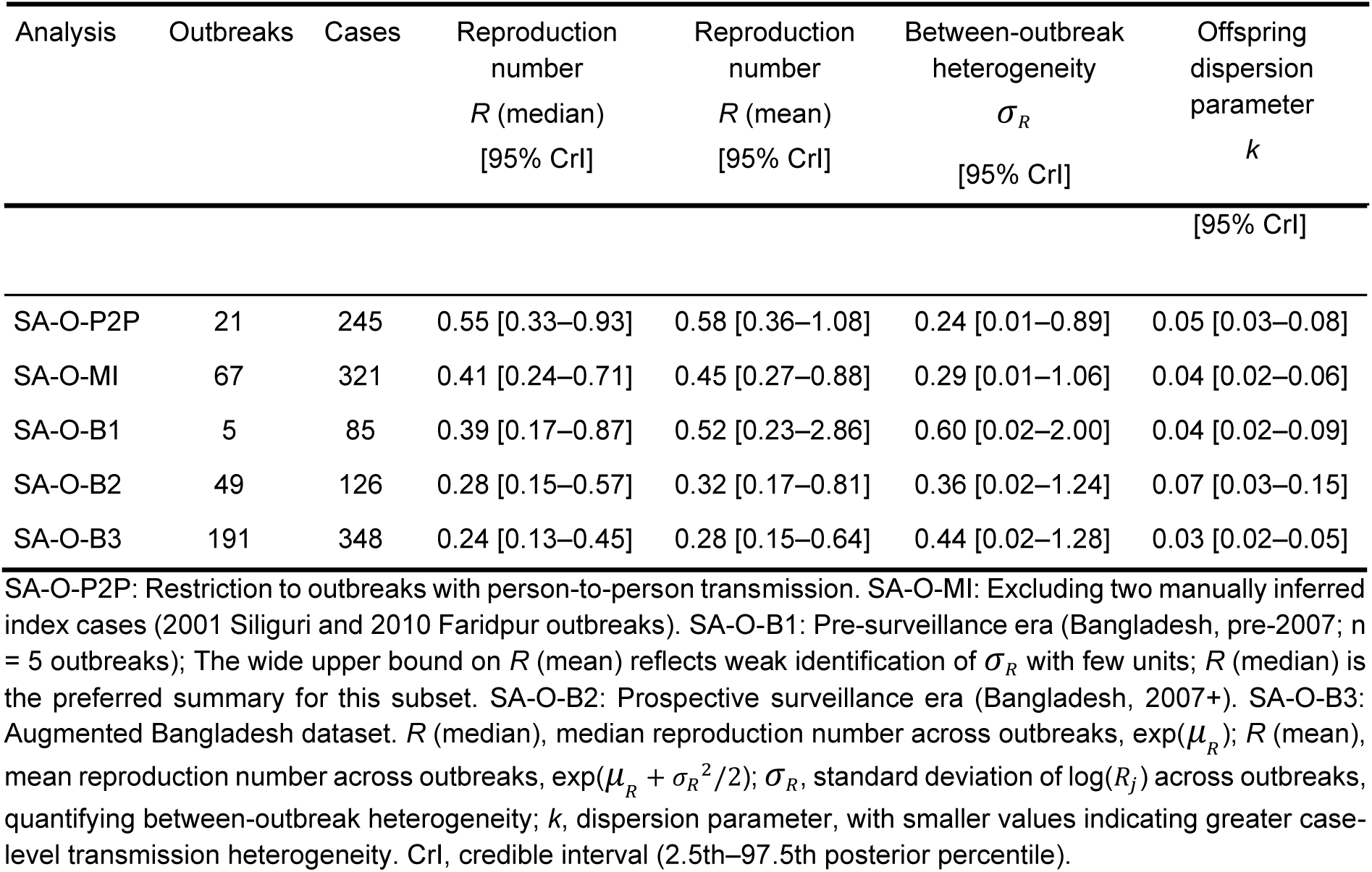
Offspring distribution sensitivity analyses.

| Analysis | Outbreaks | Cases | Reproduction number<br>$R$ (median)<br>[95% CrI] | Reproduction number<br>$R$ (mean)<br>[95% CrI] | Between-outbreak heterogeneity<br>$\sigma_R$<br>[95% CrI] | Offspring dispersion parameter<br>$k$ |
| --- | --- | --- | --- | --- | --- | --- |

| [95% CrI] |  |  |  |  |  |  |
| --- | --- | --- | --- | --- | --- | --- |
| SA-O-P2P | 21 | 245 | 0.55 [0.33–0.93] | 0.58 [0.36–1.08] | 0.24 [0.01–0.89] | 0.05 [0.03–0.08] |
| SA-O-MI | 67 | 321 | 0.41 [0.24–0.71] | 0.45 [0.27–0.88] | 0.29 [0.01–1.06] | 0.04 [0.02–0.06] |
| SA-O-B1 | 5 | 85 | 0.39 [0.17–0.87] | 0.52 [0.23–2.86] | 0.60 [0.02–2.00] | 0.04 [0.02–0.09] |
| SA-O-B2 | 49 | 126 | 0.28 [0.15–0.57] | 0.32 [0.17–0.81] | 0.36 [0.02–1.24] | 0.07 [0.03–0.15] |
| SA-O-B3 | 191 | 348 | 0.24 [0.13–0.45] | 0.28 [0.15–0.64] | 0.44 [0.02–1.28] | 0.03 [0.02–0.05] |

SA-O-P2P: Restriction to outbreaks with person-to-person transmission. SA-O-MI: Excluding two manually inferred index cases (2001 Siliguri and 2010 Faridpur outbreaks). SA-O-B1: Pre-surveillance era (Bangladesh, pre-2007; $n = 5$ outbreaks); The wide upper bound on $R$ (mean) reflects weak identification of $\sigma_R$ with few units; $R$ (median) is the preferred summary for this subset. SA-O-B2: Prospective surveillance era (Bangladesh, 2007+). SA-O-B3: Augmented Bangladesh dataset. $R$ (median), median reproduction number across outbreaks, $\exp(\mu_R)$ ; $R$ (mean), mean reproduction number across outbreaks, $\exp(\mu_R + \sigma_R^2/2)$ ; $\sigma_R$ , standard deviation of $\log(R_i)$ across outbreaks, quantifying between-outbreak heterogeneity; $k$ , dispersion parameter, with smaller values indicating greater case-level transmission heterogeneity. CrI, credible interval (2.5th–97.5th posterior percentile).

### Serial interval

The gamma distribution provided the best fit to 137 transmission pairs (ΔAIC versus lognormal = 10.0; versus Weibull = 5.1), yielding a fitted mean serial interval of 13.3 days (95% confidence interval: 12.8–13.8) and standard deviation (SD) of 3.3 days (2.7–3.9). The median of the observed pairs was 13.0 days, close to the fitted mean (Table 2). A kernel density estimate of the observed distribution is shown alongside the fitted gamma curve in Figure 5. Country-specific estimates were consistent with the pooled results: 12.8 days (12.0–13.5) in Bangladesh (n = 70 pairs) and 13.9 days (13.2–14.5) in India (n = 67 pairs). Bangladesh-specific estimate was consistent with the previously reported mean of 12.7 days (SD: 3.0) by Nikolay et al. (6). Sensitivity analyses yielded consistent estimates across all specifications (SA-S1 and SA-S2), with mean serial intervals ranging from 12.6 to 13.4 days regardless of pair inclusion criteria or surveillance era (Supplementary Table S12).

**Figure 5.**
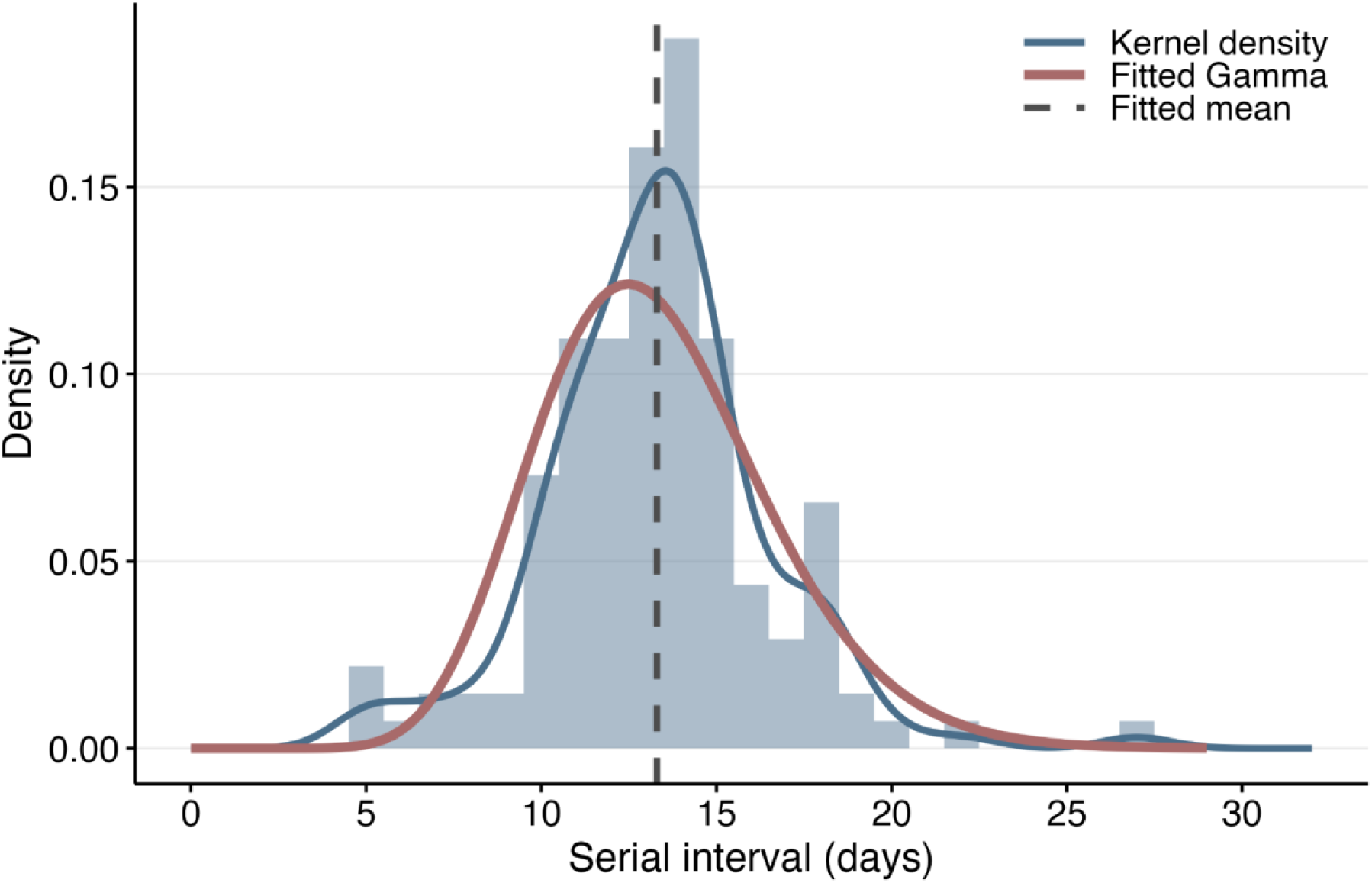
Serial interval. Distribution of observed serial intervals (n = 137 transmission pairs) with fitted gamma density curve (red) and kernel density estimate (KDE; dark blue line). The dashed vertical line indicates the fitted mean (13.3 days). The gamma distribution provided the best fit among three candidate distributions (ΔAIC versus lognormal = 10.0; versus Weibull = 5.1), with a fitted mean of 13.3 days (95% CI: 12.8–13.8) and standard deviation of 3.3 days. The KDE shows the empirical distribution is slightly more concentrated around the mean than the fitted gamma curve; the fitted and empirical standard deviations are closely matched (3.3 versus 3.2 days), suggesting the overall spread is well captured.

## Discussion

We reviewed and analysed outbreak data from Nipah virus outbreaks in Bangladesh and India spanning 2001–2026 and estimated the reproduction number, offspring dispersion, and serial interval that characterise NiV transmission. The findings suggest that the transmission is subcritical on average, with a reproduction number well below the epidemic threshold (*R* = 1); individual-level transmission is markedly overdispersed, so a minority of cases drives most onward transmission; and the mean serial interval is 13.3 days. Together, these describe transmission that is self-limiting on average but capable of stochastic amplification, in which a low reproduction number should not be equated with low risk.

The subcritical reproduction number implies that NiV transmission following zoonotic introduction can be expected to die out under current levels of transmissibility and virulence, and that sustained transmission is unlikely without repeated reintroduction. The low average reproduction number, however, should be understood alongside the dispersion parameter: most introductions die out quickly, but a minority can generate large clusters. This reconciles the two key features of NiV outbreaks, frequent dead-end spillovers and occasional larger clusters. While these transmission dynamics are consistent with prior evidence from Bangladesh (6,7,24,36), our analyses provide a formal quantitative evaluation encompassing both Bangladesh and India over a twenty-five-year period since NiV emergence in 2001 until 2026.

Bangladesh has operated a continuous, national NiV-specific hospital-based surveillance system since 2007 (6,7,63). An independent evaluation of this system estimated that approximately half of outbreaks were missed during 2007–2014, with no association found between outbreak size and distance from a surveillance hospital, the primary driver of detection in their model (63), arguing against underdetection of isolated spillovers specifically. India lacked a comparable system for most of the study period, with cases instead identified reactively following outbreaks (64), and surveillance capacity has varied within India across settings and over time (65,66). Because a substantial share of India’s person-to-person outbreaks were detected reactively, only cases severe or outbreaks large enough to be recognised may be disproportionately represented, which could inflate the degree of overdispersion in India. The available data, however, cannot distinguish a genuine country-level difference in transmission from this ascertainment effect.

Our estimated median reproduction number in Bangladesh closely matches the previous estimate published by Nikolay et al. (6). Our sensitivity analysis with augmented data for Bangladesh is similarly consistent with a recent estimate derived from surveillance data covering 2007–2018 (24), supporting the validity of both our model structure and dataset. Overdispersion in Bangladesh had previously been documented descriptively (6,24,36), and the dispersion parameter for Bangladesh has been used by Nikolay et al. 2021 and Cortes-Azuero et al. 2026 (67,68). Pan et al. conducted an analysis using a next-generation cluster approach fitted to a smaller dataset across Bangladesh and India (69), overlapping our estimate. Further, we generated comprehensively synthesised country-specific estimates of reproduction number, offspring distribution, and serial interval for India.

Our estimates carry implications for NiV prevention and control. Because transmission is self-limiting and concentrated in a minority of cases, targeted non-pharmaceutical containment such as rapid contact tracing and quarantine is expected to be more efficient than population-wide measures (70,71). Further, the reproduction number, offspring dispersion parameter, and serial interval estimated across Bangladesh and India provide empirical inputs for broader epidemiological modelling. These parameters can be leveraged to forecast outbreak trajectories, estimate the probability of nosocomial clusters, and evaluate the effectiveness of non-pharmaceutical interventions across country settings.

The reproduction number, offspring dispersion parameter, and serial interval provide key transmission parameters to assess operational constraints for deploying and evaluating medical countermeasures. While a 13.3-day serial interval, which serves as a proxy for the generation interval, outlines a temporal window for ring-based control; recent modelling studies indicate that reactive ring vaccination yields limited impact or feasibility under current NiV epidemiology (67,68,72). Additionally, as demonstrated by Hinch et al. (2026), pronounced overdispersion severely reduces the statistical power of standard cluster-randomised ring trials (73). Thus, establishing vaccine efficacy under current NiV epidemiology will likely necessitate alternative trial designs that accommodate the degree of overdispersion of NiV, or resort to regulatory pathways that do not depend on clinical disease endpoints, such as licensure based on animal model efficacy data.

Our estimates suggest that, because human transmission chains are expected to extinguish, the burden of NiV disease is driven primarily by the frequency of zoonotic spillover. Preparedness should therefore combine control of human transmission with interventions that reduce spillover at the animal-human interface, including safer date palm sap harvesting and One Health surveillance of bat populations. Because the risk is regional rather than nationally bounded, as illustrated by the concurrent appearance of cases in West Bengal and Bangladesh in January 2026 (57,62), preparedness also calls for harmonised cross-border surveillance, shared case definitions, and coordinated genomic and epidemiological data collection across the Nipah belt spanning the two countries. The estimates reported here also provide a baseline against which future shifts in transmissibility, such as through viral evolution, can be identified.

Our study has limitations. Reliance on published investigations introduces ascertainment and reporting biases, most notably the absence of outbreak-level Bangladesh data for 2015–2022. The impact of the missing cases was explored in a sensitivity analysis (SA-O-B3) that included cases with no associated outbreak record, assumed to be non-transmitting spillover introductions. Mild or asymptomatic infections might not be captured in outbreak investigations. The estimates also rest on the assumptions of the hierarchical negative binomial framework, which specifies a lognormal mixing distribution for outbreak-specific reproduction numbers and conditional independence of offspring counts given the outbreak-level parameters. The mixing distribution was not varied in the sensitivity analyses, though the small number of outbreaks and the prior-dominated between-outbreak variance make the estimates unlikely to be sensitive to this choice. Conditional independence may be approximate within outbreaks, where offspring counts are linked through the shared transmission process. These limitations are more likely to affect the precision of individual estimates and the country comparison than the central conclusions, which were consistent across prior specifications and sensitivity analyses, including exclusion of the manually inferred cases and the augmented Bangladesh analysis.

In summary, NiV in Bangladesh and India operates in a subcritical, highly overdispersed transmission regime in which individual outbreaks remain self-limiting, yet the capacity for stochastic superspreading, combined with high case-fatality and recurrent spillover, generates persistent public health risk. Further, our estimates of reproduction number, offspring dispersion, and serial interval provide the quantitative foundation for epidemiological modelling of NiV outbreaks and for assessing the potential impact of NiV vaccines in the development pipeline.

## Contributors

SK conceptualised the study, developed the methodology, curated the data, conducted the formal analysis, acquired funding, and wrote the original draft. VVM contributed to the methodology, curated and validated the data and served as the second reviewer for study screening and data extraction. JFV, HK, LS, S-mJ, and AI contributed to the methodology and reviewed and edited the manuscript. AE, WJE, and KA conceptualised the study, supervised the study, and acquired funding. All authors reviewed and approved the final manuscript.

## Data Availability

The dataset and analytical code are available at https://github.com/Sorukeem/Nipah_HNB.

## Declaration of interests

The authors have declared that no competing interests exist.

## Ethics

This study used data from published outbreak investigations and publicly available reports. Ethical approval and informed consent were not required.

## Data availability

The dataset and analytical code are available at https://github.com/Sorukeem/Nipah_HNB.

## Acknowledgments

SK, HK, LS, AE, WJE, and KA are supported by the Japan Agency for Medical Research and Development under Grant [JP223fa627004]. SK is supported by the Nagasaki University Doctoral Program for World-leading Innovative and Smart Education for Global Health, KENKYU SHIDO KEIHI. S-mJ is funded by the Singapore Ministry of Health (Programme for Research in Epidemic Preparedness and Response Co-Operative 1; PREPARE-S1-2022-02). AE is supported by the Japan Science and Technology Agency (JST) (JPMJPR22R3, JPMJFR244I) and the Japan Society for the Promotion of Science (JP22K17329). The funders had no role in study design, data collection and analysis, decision to publish, or preparation of the manuscript.

## Declaration of generative AI and AI-assisted technologies in the writing process

During the preparation of this work, the author(s) used Claude Opus 4.6 model within the Cursor editor in generating and debugging the code used for the data analysis. Following the use of this tool, the author(s) reviewed, tested, and edited the content as needed and take(s) full responsibility for the content of the publication.

## Supporting information Captions

Supplementary Texts

Supplementary Text S1. Choosing studies describing the same outbreaks

Supplementary Text S2. Analytical dataset

Supplementary Text S3. Coding decisions

Supplementary Text S4. Offspring distribution – prior specifications and sensitivity

Supplementary Text S5. Offspring distribution – sensitivity analyses

Supplementary Text S6. Results

Supplementary Text S7. Reproduction number and overdispersion parameter – existing estimates from literature

## Supplementary Tables

Supplementary Table S1. PRISMA Checklist

Supplementary Table S2. Search strategies for each database

Supplementary Table S3. Inclusion and exclusion criteria

Supplementary Table S4. List of data extracted

Supplementary Table S5. Studies included in the systematic review

Supplementary Table S6. Quality assessment results

Supplementary Table S7. Person-to-person outbreaks by transmission chain

Supplementary Table S8. Prior sensitivity

Supplementary Table S9. Country-specific analyses: P1–P3

Supplementary Table S10. Offspring distribution sensitivity analyses (SA-O-CV)

Supplementary Table S11. Derived dispersion quantities from primary model

Supplementary Table S12. Serial interval: primary and sensitivity analyses

## Supplementary Figures

Supplementary Figure S1. Epidemic curves of Nipah virus outbreaks in Bangladesh and India

Supplementary Figure S2. Prior and posterior densities under three prior specifications

Supplementary Figure S3. Posterior predictive check

Supplementary Figure S4. Observed and simulated offspring count distributions

Supplementary Figure S5. Trace plots

Supplementary Figure S6. Posterior predictive check by country

## Notes

### Competing Interest Statement

The authors have declared no competing interest.

### Author Declarations

PubMed, Embase, Web of Science, WHO Disease Outbreak News and the icddr,b Health and Science Bulletin

### Summary of Updates

Code correction, typo correction, edits for better language, etc.

