## Appendix for "Transmission dynamics of Nipah virus in Bangladesh and India, 2001–2026: systematic review and inference on reproduction number, offspring dispersion, and serial interval"

Contents

Supplementary Text S1. Choosing studies describing the same outbreaks

Supplementary Text S2. Analytical dataset

Supplementary Text S3. Coding decisions

Supplementary Text S4. Offspring distribution – prior specifications and sensitivity

1. Prior specifications

2. Prior sensitivity

Supplementary Text S5. Offspring distribution – sensitivity analyses

Supplementary Text S6. Results

1. Offspring distribution – model validation

2. Offspring distribution – country analyses

3. Offspring distribution – sensitivity analyses

Supplementary Text S7. Reproduction number and overdispersion parameter – existing estimates from literature

Supplementary Table S1. PRISMA Checklist

Supplementary Table S2. Search strategies for each database

Supplementary Table S3. Inclusion and exclusion criteria

Supplementary Table S4. List of data extracted

Supplementary Table S5. Studies included in the systematic review

Supplementary Table S6. Quality assessment results

Supplementary Table S7. Person-to-person outbreaks by transmission chain

Supplementary Table S8. Prior sensitivity

Supplementary Table S9. Country-specific analyses: P1–P3

Supplementary Table S10. Offspring distribution sensitivity analyses (SA-O-CV)

Supplementary Table S11. Derived dispersion quantities from primary model

Supplementary Table S12. Serial interval: primary and sensitivity analyses

Supplementary Figure S1. Epidemic curves of Nipah virus outbreaks in Bangladesh and India

Supplementary Figure S2. Prior and posterior densities under three prior specifications

Supplementary Figure S3. Posterior predictive check

Supplementary Figure S4. Observed and simulated offspring count distributions

Supplementary Figure S5. Trace plots

Supplementary Figure S6. Posterior predictive check by country

References

### Supplementary Text S1. Choosing studies describing the same outbreaks

1. 2001 Siliguri, India

We prioritised Harit et al. (2006) over Chadha et al. (2006) as a primary source as it presents the Nipah virus (NiV) outbreak from the outbreak investigation perspective and reports demographics, clinical presentation, and outcomes as an integrated series across the outbreak setting, while Chadha et al.'s strongest contributions are the diagnostic thresholds and molecular confirmation methods for the sampled subset, which is valuable for etiologic attribution but less directly informative for estimating time-to-event parameters at the case-series level (1,2). There are discrepancies between Harit et al. (2006) and Chadha et al. (2006) in the number of cases: 66 cases from Harit et al. (2006) and 59 from Chadha et al. (2006), stemming from the difference in the number of unconfirmed, isolated cases. Although Chadha et al. (2006) report fewer isolated cases, they provide the date of onset for those cases, which are not available from Harit et al. (2006). The infector-infectee pairs were reconstructed primarily based on Harit et al. (2006), while deriving some of the onset date data from Chadha et al. (2006). Harit et al. (2006) attribute 32 secondary cases to case 9, a patient admitted to Nursing Home 'B' on 3 February and deceased on 7 February: 24 staff of the nursing home, five attendants of case 9, and three attendants of other patients, all with probable exposure during 3–7 February and onsets between 12 and 23 February (one onset unrecorded) (2). Chadha et al. (2006) describe the same cluster as 23 staff and eight visitors, 31 secondary cases in total, following a single patient admitted to the same hospital on 3 February, with onsets between 10 and 23 February (1). The two investigations therefore agree on the structure of the cluster and on the single attributed infector and differ by one case in its size. Consistent with the prioritisation rule applied throughout, we adopted the count from the source with the more complete case ascertainment and assigned case 9 an offspring count of 32.

2. 2004 Rajbari, Bangladesh

We chose Montgomery et al. (2008) as the primary source for the infector-infectee pairs data, as it provides the most comprehensive information on outbreak investigations (3). We cross-checked the information across the icddr,b Health and Science Bulletin report (HSB) (2004) and the WHO Weekly Epidemiological Report (WER) (2004) (4,5).

3. 2004 Faridpur, Bangladesh

We chose Gurley et al. (2007) as the primary source for the infector-infectee pairs data and cross-checked it against the icddr,b HSB (2004) and the WHO WER (2004) (4–6).

4. 2005 Tangail, Bangladesh

We chose Luby et al. (2006) as the primary source for the infector-infectee pairs data and cross-checked it against the icddr,b HSB (2005) (4,7).

5. 2007 Thakurgaon, Bangladesh

We chose Homaira et al. (2010a) as the primary source for the infector-infectee pairs data and cross-checked it against the icddr,b HSB (2007) (4,8).

6. 2007 Kushtia, Bangladesh

We chose Homaira et al. (2010b) as the primary source for the infector-infectee pairs data and cross-checked it against the icddr,b HSB (2007) (4,9).

7. 2018 Kerala, India

Thulaseedaran et al. (2018), Kumar et al. (2018), Thomas et al. (2019), Chandni et al. (2020), Arunkumar et al. (2019), and Pallivalappil et al. (2020) report on the same outbreak of Kerala in 2018, which was the first outbreak in the southwestern region of India (10–13). Thulaseedaran et al. (2018), Chandni et al. (2020), and Thomas et al. (2019) are clinical case series that describe subsets of cases (10,12,13). Thus, we excluded the three studies in the full-text screening stage and included Arunkumar et al. (2019) and Pallivalappil et al. (2020) for data extraction. There are discrepancies between Arunkumar et al. (2019) and Pallivalappil et al. (2020): the age of the index case (27 from Arunkumar et al. (2019) vs. 26 from Pallivalappil et al. (2020)); Incubation period (median 9.5 days (range 6–14) vs. median 9.5 days (range 4–14)), while Pallivalappil et al. (2020) has internal inconsistencies around incubation period values (Table 1 shows 10.5 days); and contact tracing: 2642 contacts vs. 2649 contacts. We prioritised Arunkumar et al. (2019) as the data source for this outbreak, as they provide the most complete details on infector-infectee pairs (14).

8. 2011 Lalmonirhat, Bangladesh

While Chakraborty et al. (2016) identify 22 cases with person-to-person transmission, icddr,b HSB (2011) reports 20 cases with no person-to-person transmission (4,15). The newly identified case was a janitorial staff member who was a probable case without a specimen collected (15). This was the second healthcare worker infected with NiV in a nosocomial setting in Bangladesh (15). We chose Chakraborty et al. (2016) as the source of the infector-infectee transmission pair reconstruction (15).

9. 2011 Rangpur, Bangladesh

While Chakraborty et al. (2016) capture eight cases, icddr,b HSB (2012) reports seven cases (4,15). Considering right truncation, we include the last case identified by Chakraborty et al. (2016) and include this case in the infector-infectee pairs. At the same time, icddr,b HSB (2012) provides details on the date of onset, while Chakraborty et al. (2016) do not. Islam et al. (2016) also report the 2011 Rangpur outbreak, but with seven cases (16). Thus, we excluded Islam et al. (2016) for this specific outbreak and included Chakraborty et al. (2016) and icddr,b HSB (2012).

10. 2023 Rajbari, Bangladesh

We prioritised Rahman et al. (2025) over the WHO Disease Outbreak News (2023) for the 2023 Rajbari outbreak as Rahman et al. (2025) further identified a secondary case which was not reported in the WHO DONs (2023) (17,18).

11. 2025 West Bengal, India

We used WHO DONs (2026) as the primary source for infector-infectee pairs data and derived outcome-related information from the WHO SEARO report (2026) (19,20).

### Supplementary Text S2. Analytical dataset

We constructed a case-level transmission pair dataset from the included studies. Each row represents an infector-infectee pair. The analytical dataset comprised 323 cases across 67 outbreaks in Bangladesh (n = 211 cases, 54 outbreaks) and India (n = 112 cases, 13 outbreaks). For each human case, defined as any individual appearing as an infectee in the dataset, we derived the offspring count $X_{ij}$ as the number of person-to-person transmission pairs in which that individual subsequently appeared as an infector. Cases that never appeared as an infector were assigned $X_{ij} = 0$. Non-human, environmental, or unidentified sources appearing only as infectors, not as infectees, such as bats or contaminated date palm sap, were excluded from the offspring distribution denominator. Their downstream infectees were retained as cases and contributed $X_{ij} = 0$ if they generated no further transmission.

### Supplementary Text S3. Coding decisions

Some outbreak-specific coding decisions warrant documentation. Case count for the 2001 Siliguri outbreak increased by 1 (66 from Harit et al. (2006); 67 in the dataset) (2). The primary case, though unidentified, has a precisely attributed offspring count of 9 from Harit's Figure 2 transmission diagram and text description, but was not counted as a case in the source study. Excluding this case would discard a valid observation of an unusually high offspring count, biasing the offspring distribution toward underdispersion. We therefore include this case as an infector with an offspring count of 9, with an unknown onset date, and an unknown transmission setting (coded accordingly).

In the 2001 Meherpur (Bangladesh) outbreak, 11 secondary cases (A1-A4, B1-B2, S1-S5) were attributed to one infectee case as their most likely infector but with residual uncertainty about whether transmission was direct or via a common household source, as acknowledged by the original investigation (21).

Case count for the 2010 Faridpur (Bangladesh) outbreak increased by 1 (16 from Sazzad et al. (2013); 17 in the dataset) (22). The article suggested the possibility of a common infector for the two confirmed cases (one admitted patient with no history of date palm sap consumption and one physician). An unidentified NiV-infected case-patient in the same adult male ward, given the context, is considered highly plausible and included in the dataset as an unknown primary case.

The 2011 Rajbari (Bangladesh) outbreak with two cases was reported by Chakraborty et al. (2016) (15). The first case (mother, deceased) and second case (daughter, deceased) had both contact history after the onset of illness of the first case (4 days prior to the second case's onset of illness) and date palm sap consumption history before their onset of illness (within 3 weeks). It is not clear whether they consumed the same contaminated date palm sap. We code these two cases as a transmission pair, not as two isolated cases.

When there were multiple candidate infectors (number of infectees with multiple candidate infectors in our dataset = 4), we applied a set of rules to choose the one infector for the negative binomial model in order: the earliest recorded exposure start; the earliest infector symptom onset; whether the infector died. If more than one candidate remained, we drew one at random after sorting infector identifiers. In the four affected infectees, no candidate was linked to any other infectee, so the offspring count distribution is identical under any assignment, and the choice determines only which individual carries the count. Serial interval pairs from these cases were included in the primary serial interval analysis but flagged for the sensitivity analysis restricting to unambiguous transmission pairs (SA-S1).

### Supplementary Text S4. Offspring distribution – prior specifications and sensitivity

#### 1. *Prior specifications*

(1) P1 (weakly informative reference): $\mu_R$ ~ Normal(0, 1); $\sigma_R$ ~ HalfNormal(0, 1); $k$ ~ Exponential(1). P1 imposes no pathogen-specific information and serves as a reference to assess whether $R$ and $k$ are data-driven or prior-dependent. The Exponential(1) prior on $k$ has median 0.69 and places approximately 4% of its mass below 0.04, so it disfavours the range in which the posterior falls; agreement between P1 and the informative specifications therefore constitutes a stringent test of prior-dependence for $k$.

(2) P2 (epidemiologically informed, permissive heterogeneity): $\mu_R$ ~ Normal(log(0.33), 0.5); $\sigma_R$ ~ HalfNormal(0, 1); $k$ ~ LogNormal(-2.5, 0.8). The prior on $\mu_R$ is centred at a prior median $R$ of 0.33, consistent with the existing Bangladesh estimate (Nikolay et al.: 0.33 [0.19–0.59]) (23), with 95% prior mass spanning approximately [0.12, 0.90]. The prior on $k$ has median exp(-2.5) ≈ 0.08, of the same order as the value of $k$ = 0.06 for Bangladesh used by Nikolay et al. (24). The permissive HalfNormal(0, 1) prior on $\sigma_R$ allows between-outbreak heterogeneity to be large or small without constraint.

(3) P3 (epidemiologically informed, conservative heterogeneity): priors on $\mu_R$ and $k$ as in P2; $\sigma_R$ ~ HalfNormal(0, 0.25). The constrained $\sigma_R$ prior concentrates 95% of prior mass below 0.49, testing whether conclusions are sensitive to the assumption of low between-outbreak heterogeneity.

### 2. *Prior sensitivity*

Posterior estimates of $R$ and $k$ were consistent across all three prior specifications (Table S8). Median $R$ ranged from 0.43 (95% CrI: 0.26–0.75) under P2 to 0.52 (0.29–1.04) under P1, with P3 giving 0.44 (0.27–0.74). The posterior mass for $R$ lay predominantly below 1 under all three specifications; under the weakly informative P1, the upper bound of the credible interval marginally exceeded unity (1.04 for median $R$; 1.24 for mean $R$), whereas under both epidemiologically informed specifications (P2 and P3) the entire interval lay below 1. The dispersion parameter $k$ was invariant across specifications at 0.04 (P1: 0.02–0.06; P2 and P3: 0.03–0.06), indicating that the posterior for $k$ is determined by the data rather than by the prior specifications examined (Figure S2).

The between-outbreak heterogeneity parameter $\sigma_R$ was weakly identified. Under P1 and P2, which share a HalfNormal(0, 1) prior, the posterior shifted downward relative to the prior—posterior medians of 0.27 and 0.28 in the pooled analysis against a prior median of 0.674—but remained comparably diffuse across orders of magnitude (Figure S2). The shift was largest in the pooled analysis (67 outbreaks) and smallest in the India analysis (13 outbreaks), as expected for a variance component informed by the number of outbreak-level units rather than by the number of cases. The data therefore locate $\sigma_R$ only loosely: they disfavour the largest values admitted by the prior but do not distinguish small positive values from zero. Under P3, the posterior was indistinguishable from the prior in all three analyses (posterior medians 0.15–0.17 against a prior median of 0.169; Figure S2), so the narrower $\sigma_R$ posterior under P3 reflects the constrained HalfNormal(0, 0.25) prior rather than greater data informativeness. Because P1 and P2 impose the same prior on $\sigma_R$, the difference between their $\sigma_R$ posteriors is not informative about prior sensitivity for this parameter; only the P3 contrast is, and it represents a prior assumption rather than an empirical finding.

P2 was used as the primary specification for all reported estimates. $R$ and $k$ are robust across specifications, so this choice does not materially affect those estimates; it matters principally for $\sigma_R$ and for the derived mean $R$. P2 was preferred over P1 because its priors on $\mu_R$ and $k$ encode the available epidemiological information on NiV transmission in Bangladesh rather than discarding it, while remaining diffuse enough to be updated by the data, and over P3 because its permissive $\sigma_R$ prior does not impose a degree of between-outbreak heterogeneity that the data cannot support in either direction.

*MCMC settings and convergence.* Four chains were run with 1,000 warm-up and 2,000 sampling iterations (8,000 post-warm-up draws), with a target acceptance rate of 0.95, and a maximum tree depth of 12. All three specifications achieved satisfactory MCMC convergence, with maximum $\hat{R}$ < 1.003 (target < 1.01), minimum bulk effective sample sizes > 2,500 (target > 400), and zero divergent transitions.

### Supplementary Text S5. Offspring distribution – sensitivity analyses

SA-O-P2P: Person-to-person-confirmed outbreaks only. We restricted the denominator to outbreaks in which at least one person-to-person transmission pair was documented, excluding the spillover-only outbreaks. This analysis conditions on observing transmission and is therefore expected to yield upwardly biased $R$. It is reported as an approximate upper bound and for comparability with earlier studies that applied a similar restriction (25,26).

SA-O-MI: Manually inferred cases excluded. Two index cases were manually inferred and included in the primary dataset based on strong circumstantial evidence rather than direct enumeration in the source studies (Text S3): an unidentified primary case in the 2001 Siliguri outbreak, assigned an offspring count of 9 (2); and an unidentified primary case in the 2010 Faridpur outbreak, assigned an offspring count of 2 (22). To assess whether these two reconstructions were influential on the offspring dispersion estimate, both cases were excluded from the pooled dataset and the primary model refit under prior set P2.

SA-O-B1: Pre-surveillance era. In Bangladesh, hospital-based surveillance and standardised contact investigations were implemented in 2007 (23,27). In the SA-O-B1 sensitivity analysis, we restricted the Bangladesh data to outbreaks from the pre-surveillance era, covering the period from 2001 to 2006.

SA-O-B2: Prospective surveillance era. We restricted the Bangladesh data to outbreaks from 2007 onwards, with hospital-based surveillance, and standardised contact investigations were implemented. All India outbreaks were excluded in SA-O-B1 and SA-O-B2.

SA-O-B3: Augmented Bangladesh analysis. To explore the impact of missing cases without outbreak records on the offspring distribution parameters, we constructed an augmented Bangladesh dataset incorporating 137 cases in addition to those retained in the systematic review. The Institute of Epidemiology, Disease Control and Research (IEDCR) reports the number of cases by year and by district, covering 2001 to 2026 at https://iedcr.gov.bd/ (Total number of cases = 348) (28). The current dataset established through the systematic review includes 211 Bangladesh cases. The difference (n = 137) was assumed to represent cases from outbreaks not identified through the review's search strategy or not meeting inclusion criteria, predominantly attributable to the 2015–2022 reporting gap described in the main text and was assumed to consist of non-transmitting spillover introductions. Each of the 137 cases was therefore added as an independent single-case outbreak with an offspring count of zero; this assumption could not be verified at the individual-case level and represents a simplification, since some of these cases may in principle have arisen from unrecognised transmission chains. We fit the augmented dataset (191 outbreaks, 348 cases) under prior P2 using the same hierarchical structure, restricted to Bangladesh.

<u>SA-O-CV</u>: Fixed variance-to-mean ratio (reparameterised dispersion). The primary model assumes a single offspring dispersion parameter $k$ shared across all cases and outbreaks, with outbreak-specific reproduction numbers $R_j$ drawn from a lognormal mixing distribution. Under this specification, the variance of the offspring distribution for an outbreak with reproduction number $R_j$ is $Var(X_{ij}) = R_j + R_j^2/k$, so the variance-to-mean ratio (VMR) equals $1 + R_j/k$ and increases with $R_j$. Consequently, outbreaks with higher reproduction numbers are implicitly assigned a higher variance-to-mean ratio, even though $k$ is shared. SA-O-CV instead constrains the variance-to-mean ratio to be constant across all outbreaks, so that the dispersion parameter varies with $R_j$ rather than being shared, and examines how the estimates change under this alternative specification.

*Model specification*. SA-O-CV model retains the hierarchical lognormal mixing distribution for outbreak-specific reproduction numbers unchanged:

$$log(R_j) \sim Normal(\mu_R, {\sigma_R}^2)$$

The shared parameter is now $\psi = R_j/k_j$ (the $R/k$ ratio), which is held constant across all outbreaks. The outbreak-specific dispersion parameter is derived as:

$$k_j = R_j/\psi$$

so that the number of secondary cases for case $i$ in outbreak $j$ follows:

$$X_{ij}|R_j, \psi \sim NegBin(R_j, k_j = R_j/\psi)$$

Under this parameterisation, $Var(X_{ij}) = R_j(1 + \psi)$ and VMR = $1 + \psi$, which is constant regardless of $R_j$. Holding $\psi$ constant therefore makes the variance proportional to the mean across outbreak, in contrast to the primary model, in which $k$ is shared and the variance grows faster than the mean as $R_j$ increases. As $\psi \rightarrow 0$, $k_j \rightarrow \infty$ and the distribution converges to Poisson; as $\psi \rightarrow \infty$, the variance-to-mean ratio grows without bound.

The coefficient of variation of the offspring distribution at outbreak-specific reproduction number $R_j$ is CV=$\sqrt{}(1/R_j + 1/k_j) = \sqrt{}((1 + \psi)/R_j)$, which decreases with $R_j$, as it does under the primary parameterisation (CV=$\sqrt{}(1/R_j + 1/k)$). The implied dispersion at the median reproduction number, $k$ at the median $R$ = $R_{med}/\psi$ with $R_{med}$ = exp($\mu_R$), is reported to allow direct comparison with the primary model's shared $k$.

*Prior specification.* Priors on $\mu_R$ and $\sigma_R$ are identical to P2 in the primary analysis: $\mu_R \sim Normal(log(0.33), 0.5)$ and $\sigma_R \sim HalfNormal(0, 1)$. The prior for $\psi$ is $\psi \sim LogNormal(1.39, 0.8)$: with $k \sim LogNormal(-2.5, 0.8)$, the reciprocal 1/$k$ is $LogNormal(2.5, 0.8)$, and multiplying by $R_j$ = 0.33 gives $\psi \sim LogNormal(2.5 + log0.33, 0.8) = LogNormal(1.39, 0.8)$, with median 4.0 (VMR = 1 + $\psi$ = 5.0) and 95% prior mass spanning approximately [0.84, 19.3].

*MCMC settings.* Four chains, 1,000 warm-up iterations, 2,000 sampling iterations per chain (8,000 post-warm-up draws). Target acceptance rate 0.95, maximum tree depth 12. Convergence assessed by $\hat{R} < 1.01$ and bulk effective sample size, identical to the primary analysis.
*Implementation.* The model was implemented in Stan via the cmdstanr interface, with a non-centred parameterisation for the outbreak-specific random effects, identical to the primary model (29–31). $\psi$ is the sole estimated dispersion quantity; the outbreak-specific dispersion parameters $k_j = R_j/\psi$ and the summary quantity k_at_R$_{med}$ = $R_{med}/\psi$ are computed in the generated quantities block for each posterior draw and summarised as posterior medians with 95% credible intervals.
*Rationale.* The two specifications differ in how dispersion is held constant across outbreaks. Under a shared $k$, the gamma mixing distribution of individual transmissibility, in the gamma-Poisson representation of the negative binomial, has the same shape parameter in every outbreak, and the variance-to-mean ratio of offspring counts grows with $R_j$. Under a shared $\psi$, the variance-to-mean ratio is constant and the shape parameter varies with $R_j$. The two parameterisations are not nested and imply different scaling of transmission heterogeneity with the reproduction number.

### Supplementary Text S6. Results

#### 1. Offspring distribution – model validation

Model adequacy was assessed by comparing three summary statistics from the observed data to 1,000 posterior predictive replicates under the P2 prior (Figure S3). Statistics were: the mean offspring count per case (average transmission intensity); the proportion of cases responsible for 80% of onward transmission (P80; superspreading concentration); and the proportion of outbreaks in which all cases generated zero secondary infections (outbreak-level self-limitation). The third statistic operates at the outbreak level rather than the case level, complementing the two case-level statistics by operating at the outbreak level. Two-sided Monte Carlo *p*-values were computed; values near 1 indicate the observed statistic is well-centred within the predictive distribution, and values near 0 indicate a poor fit.

All three observed statistics fell within the 95% predictive intervals (MC *p* 0.30–1.00). The mean offspring count (observed 0.49 secondary cases per case; MC *p* = 0.92) suggested the model reproduces the average transmission intensity. P80 (observed 4.0% of cases responsible for 80% of transmission; MC *p* = 1.00) summarises the concentration of transmission in the fitted data. The outbreak-level proportion of self-limiting outbreaks, defined as outbreaks in which no case generated a secondary infection, was 68.7% in the observed data (46 of 67 outbreaks) against a predictive median of 74.6% (50 of 67) and a 95% predictive interval of 65.6–83.6% (MC *p* = 0.30). The observed value lies inside the interval but below the predictive median, so this check is adequate rather than tightly centred. Visual comparison of the observed and marginal predicted offspring distributions showed close concordance, especially at x = 0 (~90% of cases); the largest discrete discrepancy was a modest excess of observed counts at x = 4 (Figure S4).

Between-outbreak heterogeneity parameter $\sigma_R$ is not directly checkable by these posterior predictive statistics. Because the replicates are generated conditional on the fitted outbreak-specific reproduction numbers, any between-outbreak variance statistic is reproduced by construction, and these checks are therefore not informative about $\sigma_R$. This is a data limitation rather than a modelling failure and is acknowledged in the study limitations.

Country-specific posterior predictive checks showed adequate fit in both Bangladesh and India (Figure S6). MC *p*-values ranged from 0.57 to 1.00 in Bangladesh and from 0.42 to 1.00 in India. Wider predictive intervals for India reflect the smaller sample size (13 outbreaks, 112 cases).

### 2. *Offspring distribution – country analyses*

The median $R$ was 0.47 (95% CrI: 0.22–0.98) for India and 0.35 (0.19–0.63) for Bangladesh under prior set P2 (Table S9). India's credible interval was wide, and its upper bound approached 1. The between-country contrast was inconclusive: the posterior probability that $R$ was higher in India than Bangladesh ($R_{India} > R_{Bangladesh}$) was 0.73, with a median India-to-Bangladesh ratio of 1.33 (95% CrI: 0.51–3.42), the ratio interval spanning unity.

The dispersion parameter $k$ was 0.03 (0.02–0.06) for India and 0.05 (0.03–0.09) for Bangladesh (Table S9). The posterior probability that $k$ was lower in India than Bangladesh ($k_{India} < k_{Bangladesh}$) was 0.85 (median ratio 0.65 [0.26–1.48]). Both countries showed high overdispersion ($k \ll 1$), indicating that pronounced individual-level heterogeneity is not specific to the Bangladesh setting.

The Indian reproduction number was sensitive to prior specification, reflecting the small sample (13 outbreaks, 112 cases). Under the weakly informative prior (P1), the Indian median $R$ was 0.84 (0.30–3.04), spanning subcritical to supercritical values; under the epidemiologically informed prior (P2), it was 0.47 (0.22–0.98) (Table S9). The dispersion parameter $k$ was stable across prior specifications. Under P3, the $\sigma_R$ posterior was narrower than under P1 and P2 in both the country-specific and pooled analyses, but reproduced the constrained HalfNormal(0, 0.25) prior rather than reflecting greater data informativeness (Figure S2). Median $R$ and $k$ shifted little under this constraint.

A further stratification by genotype (NiV-B, NiV-I) was considered, but the NiV-I data (Kerala outbreaks) were even smaller (10 outbreaks, 37 cases with 7 transmitters). Country was therefore retained as the stratifying variable. The country-specific estimates provide setting-specific inputs relevant to national or regional preparedness and response planning, and can be updated as further outbreak data accumulate.

#### 3. *Offspring distribution – sensitivity analyses*

Subcritical transmission and high overdispersion were suggested across all sensitivity specifications (Table 3, Table S10). Restriction to person-to-person outbreaks (SA-O-P2P) yielded $R$ = 0.55 (95% CrI: 0.33–0.93) and $k$ = 0.05 (0.03–0.08). Excluding spillover-only outbreaks removes cases whose zero offspring counts are absorbed by near-zero outbreak-specific $R_j$ rather than by the shared dispersion parameter; the restriction therefore raises $R$ substantially while leaving $k$ largely unchanged, with the implied $R/k$ essentially invariant. The SA-O-P2P $R$ estimate is consistent with prior estimates from similarly restricted analyses (Luby et al. 2009: $R$ = 0.48; Naser et al. 2015: $R$ = 0.75–0.88) (25,26) and is reported as an approximate upper bound on $R$.

Excluding the two manually inferred index cases (SA-O-MI) yielded $R$ = 0.41 (0.24–0.71) and $k$ = 0.04 (0.02–0.06) (67 outbreaks, 321 cases), showing little impact on the offspring dispersion estimates (Table 3).

Bangladesh surveillance-stratification yielded median $R$ = 0.39 (0.17–0.87) in the pre-surveillance period (SA-O-B1; 2001–2006; 5 outbreaks, 85 cases) and 0.28 (0.15–0.57) under prospective surveillance from 2007 onward (SA-O-B2; 2007–2026; 49 outbreaks, 126 cases); the credible intervals overlap substantially, and the pre-surveillance estimate carries considerable uncertainty given the limited number of outbreaks in that era. The SA-O-B2 estimate is consistent with the similar constraints investigated in Nikolay et al. 2019 (cases identified during 2007–2014: $R$ = 0.23 [95% CI 0.11–0.46]) (23).

The augmented Bangladesh analysis (SA-O-B3), which added 137 cases without outbreak-level information as presumed non-transmitting single-case spillovers, yielded $R$ = 0.24 (0.13–0.45) and $k$ = 0.03 (0.02–0.05) (191 outbreaks, 348 cases), compared with $R$ = 0.35 (0.19–0.63), $\sigma_R$ = 0.39 (0.02–1.24), and $k$ = 0.05 (0.03–0.09) in the Bangladesh country analysis (Table 2). Because $R$, $\sigma_R$ and $k$ are jointly estimated within a Bayesian hierarchical model fitted to the same offspring data under prior set P2, the coordinated decrease in both $R$ and $k$ reflects an update to the joint posterior as a whole, and a direct parameter-by-parameter comparison against the country-specific estimates is not straightforward. Unlike SA-O-P2P, where the restriction and its expected direction of bias are well characterised, the number, size, and transmission history of the 137 added cases cannot be verified, and the supplemented estimates should be interpreted as illustrative of the plausible influence of missing cases rather than as a corrected estimate. Because every supplemented case is assigned a zero offspring count by construction, this specification is conservative with respect to $R$ but not with respect to $k$: it can only increase the estimated degree of overdispersion. The qualitative conclusions of subcritical transmission and substantial overdispersion were retained under this sensitivity analysis (Table 3).

SA-O-CV (fixed variance-to-mean ratio) yielded $R$ (median) = 0.40 (0.25–0.66), $\psi$ = 10.22 (5.90–19.08), implied $k$ at the median $R$ of 0.04 (0.02–0.06), and $\sigma_R$ = 0.24 (0.01–0.72) (Table S10). The entire 95% CrI for $R$ lies below 1, suggesting subcritical transmission under this alternative dispersion structure. Internal consistency was suggested by the close agreement between $\psi$ fitted under SA-O-CV (10.22 [5.90–19.08]) and $\psi$ derived from the primary model posterior without refitting (10.87 [5.72–22.00]; Table S11), with substantially overlapping credible intervals. The implied $k$ at the median $R$ (0.04 [0.02–0.06]) was likewise consistent with the primary shared $k$ (0.04 [0.03–0.06]), suggesting that high overdispersion is robust to the parametric form of the dispersion structure. MCMC diagnostics were satisfactory: maximum $\hat{R}$ = 1.002, minimum bulk ESS > 2,500, zero divergent transitions.

### Supplementary Text S7. Reproduction number and overdispersion parameter – existing estimates from literature

Published transmission parameter estimates for NiV derive almost exclusively from Bangladesh. Luby et al. 2009 reported reproduction number of 0.48 for cases identified during 2001–2007 (26), and Naser et al. 2015 reported reproduction number of 0.75–0.88 from cluster- and case-based surveillance (25); both were derived from restricted case sets and are comparable to our person-to-person restricted analysis (SA-O-P2P: $R$ = 0.55 [0.33–0.93]). Nikolay et al. 2019 reported a reproduction number of 0.33 (95% CI: 0.19–0.59) across 2001–2014, with subset estimates of 0.23 (95% CI: 0.11–0.46) for cases identified during 2007–2014, 0.30 for primary cases, and 0.60 for cases identified late or not hospitalised (23). Nikolay et al. 2020 subsequently reported a reproduction number of 0.2 (95% CI: 0.1–0.4) from surveillance data covering 2007–2018 (32).

Our Bangladesh estimate (median $R$ = 0.35 [95% CrI: 0.19–0.63]) lies within this range and is closest to the Nikolay et al. 2019 estimate. Prior set P2 centres $\mu_R$ at log(0.33), taken from that study (Text S4). The weakly informative specification P1 yields (0.42 [0.21–0.94]; Table S9). Our surveillance-era estimates are likewise consistent with the surveillance-era literature (SA-O-B2: 0.28 [0.15–0.57] versus 0.23; SA-O-B3: 0.24 [0.13–0.45] versus 0.20). These are consistency checks against independently assembled datasets, not validation; SA-O-B3 additionally rests on assumptions about the 137 added cases (Text S5).

Overdispersion in Bangladesh had previously been documented descriptively (8% of cases responsible for onward transmission) (6,23,32). Nikolay et al. 2019 reported that a small minority of cases accounted for the majority of onward transmission (23), and the largest number of secondary cases attributed to a single infector has been reported as 21 (Nikolay et al. 2019) and 22 (Gurley et al. 2007), although a recent independent systematic review of NiV epidemiological parameters was unable to verify whether these refer to the same individual (6,23,33).

A dispersion parameter of $k$ = 0.06 has been used as a model input for Bangladesh by Nikolay et al. 2021 (24), where it reported in supplementary material with reference to Nikolay et al. 2020 (32), in which a numeric $k$ is not found, and subsequently by Cortes-Azuero et al. 2026 (34). The two studies pair it with different reproduction numbers (0.20 and 0.33, respectively). Our P2 prior (median $k \approx 0.08$) is of the same order but not centred on it (Text S4). Pan et al. 2026 reported $k$ = 0.03 (95% CrI: 0.02–0.06) using a next-generation cluster approach fitted to 32 next-generation cluster observations across Bangladesh (25 clusters) and India (7 clusters) (35); this overlaps our pooled estimate ($k$ = 0.04 [0.03–0.06]) and both country-specific estimates (Bangladesh 0.05 [0.03–0.09]; India 0.03 [0.02–0.06]), despite the difference in observational unit and dataset size.

No country-specific estimate of either parameter has previously been published for India. Our India estimates therefore have no direct comparator, and the reproduction number in particular was sensitive to prior specification (0.47 [0.22–0.98] under P2 versus 0.84 [0.30–3.04] under P1; Table S9), reflecting the limited number of outbreaks (13 outbreaks, 112 cases) rather than genuine parameter instability. The dispersion parameter was stable across prior specifications in both countries. The estimates reported here are intended as a reproducible baseline that can be updated as further outbreak data accumulate.

### Supplementary Table S1. PRISMA Checklist

| Section and Topic | Item # | Checklist item | Location where item is reported |
|---|---|---|---|
| **TITLE** | | | |
| Title | 1 | Identify the report as a systematic review. | Title |
| **ABSTRACT** | | | |
| Abstract | 2 | See the PRISMA 2020 for Abstracts checklist. | Abstract |
| **INTRODUCTION** | | | |
| Rationale | 3 | Describe the rationale for the review in the context of existing knowledge. | Introduction |
| Objectives | 4 | Provide an explicit statement of the objective(s) or question(s) the review addresses. | Introduction |
| **METHODS** | | | |
| Eligibility criteria | 5 | Specify the inclusion and exclusion criteria for the review and how studies were grouped for the syntheses. | Supplementary Table S3 |
| Information sources | 6 | Specify all databases, registers, websites, organisations, reference lists and other sources searched or consulted to identify studies. Specify the date when each source was last searched or consulted. | Methods – Search strategy and study selection; Table S2 |
| Search strategy | 7 | Present the full search strategies for all databases, registers and websites, including any filters and limits used. | Supplementary Table S2 |
| Selection process | 8 | Specify the methods used to decide whether a study met the inclusion criteria of the review, including how many reviewers screened each record and each report retrieved, whether they worked independently, and if applicable, details of automation tools used in the process. | Methods – Search strategy and study selection |
| Data collection process | 9 | Specify the methods used to collect data from reports, including how many reviewers collected data from each report, whether they worked independently, any processes for obtaining or confirming data from study investigators, and if applicable, details of automation tools used in the process. | Methods – Search strategy and study selection |
| Data items | 10a | List and define all outcomes for which data were sought. Specify whether all results that were compatible with each outcome domain in each study were sought (e.g. for all measures, time points, analyses), and if not, the methods used to decide which results to collect. | Methods – Search strategy and study selection; Table S4 |
| | 10b | List and define all other variables for which data were sought (e.g. participant and intervention characteristics, funding sources). Describe any assumptions made about any missing or unclear information. | Table S4; Text S1 |

| | | | |
|---|---|---|---|
| Study risk of bias assessment | 11 | Specify the methods used to assess risk of bias in the included studies, including details of the tool(s) used, how many reviewers assessed each study and whether they worked independently, and if applicable, details of automation tools used in the process. | Methods – Quality assessment |
| Effect measures | 12 | Specify for each outcome the effect measure(s) (e.g. risk ratio, mean difference) used in the synthesis or presentation of results. | N/A |
| Synthesis methods | 13a | Describe the processes used to decide which studies were eligible for each synthesis (e.g. tabulating the study intervention characteristics and comparing against the planned groups for each synthesis (item #5)). | Text S1 |
| | 13b | Describe any methods required to prepare the data for presentation or synthesis, such as handling of missing summary statistics, or data conversions. | Methods – Transmission pair reconstruction |
| | 13c | Describe any methods used to tabulate or visually display results of individual studies and syntheses. | Text S2 |
| | 13d | Describe any methods used to synthesize results and provide a rationale for the choice(s). If meta-analysis was performed, describe the model(s), method(s) to identify the presence and extent of statistical heterogeneity, and software package(s) used. | Methods – Offspring distribution model; Serial interval estimation |
| | 13e | Describe any methods used to explore possible causes of heterogeneity among study results (e.g. subgroup analysis, meta-regression). | Methods – Offspring distribution model; Serial interval estimation |
| | 13f | Describe any sensitivity analyses conducted to assess robustness of the synthesized results. | Methods – Offspring distribution model; Serial interval estimation; Text S5 |
| Reporting bias assessment | 14 | Describe any methods used to assess risk of bias due to missing results in a synthesis (arising from reporting biases). | Methods – Quality assessment |
| Certainty assessment | 15 | Describe any methods used to assess certainty (or confidence) in the body of evidence for an outcome. | Methods – Offspring distribution model; Serial interval estimation |
| **RESULTS** | | | |
| Study selection | 16a | Describe the results of the search and selection process, from the number of records identified in the search to the number of studies included in the review, ideally using a flow diagram. | Results – Study selection and quality assessment; Figure 1 |
| | 16b | Cite studies that might appear to meet the inclusion criteria, but which were excluded, and explain why they were excluded. | Text S1 |
| Study characteristics | 17 | Cite each included study and present its characteristics. | Table S5 |
| Risk of bias in studies | 18 | Present assessments of risk of bias for each included study. | Table S6 |

| | | | |
|---|---|---|---|
| Results of individual studies | 19 | For all outcomes, present, for each study: (a) summary statistics for each group (where appropriate) and (b) an effect estimate and its precision (e.g. confidence/credible interval), ideally using structured tables or plots. | Table 1 |
| Results of syntheses | 20a | For each synthesis, briefly summarise the characteristics and risk of bias among contributing studies. | Results – Offspring distribution; Serial interval |
| | 20b | Present results of all statistical syntheses conducted. If meta-analysis was done, present for each the summary estimate and its precision (e.g. confidence/credible interval) and measures of statistical heterogeneity. If comparing groups, describe the direction of the effect. | Results – Offspring distribution; Serial interval |
| | 20c | Present results of all investigations of possible causes of heterogeneity among study results. | Results – Offspring distribution; Serial interval; Text S6 |
| | 20d | Present results of all sensitivity analyses conducted to assess the robustness of the synthesized results. | Results – Offspring distribution; Serial interval; Text S6 |
| Reporting biases | 21 | Present assessments of risk of bias due to missing results (arising from reporting biases) for each synthesis assessed. | Results – Offspring distribution; Serial interval; Discussion |
| Certainty of evidence | 22 | Present assessments of certainty (or confidence) in the body of evidence for each outcome assessed. | Results – Offspring distribution; Serial interval |
| **DISCUSSION** | | | |
| Discussion | 23a | Provide a general interpretation of the results in the context of other evidence. | Discussion |
| | 23b | Discuss any limitations of the evidence included in the review. | Discussion |
| | 23c | Discuss any limitations of the review processes used. | Discussion |
| | 23d | Discuss implications of the results for practice, policy, and future research. | Discussion |
| **OTHER INFORMATION** | | | |
| Registration and protocol | 24a | Provide registration information for the review, including register name and registration number, or state that the review was not registered. | Methods – Search strategy and study selection |
| | 24b | Indicate where the review protocol can be accessed, or state that a protocol was not prepared. | Methods – Search strategy and study selection |
| | 24c | Describe and explain any amendments to information provided at registration or in the protocol. | N/A |
| Support | 25 | Describe sources of financial or non-financial support for the review, and the role of the funders or sponsors in the review. | Acknowledgements |
| Competing interests | 26 | Declare any competing interests of review authors. | Declaration of interests |

| | | | |
|---|---|---|---|
| Availability of data, code and other materials | 27 | Report which of the following are publicly available and where they can be found: template data collection forms; data extracted from included studies; data used for all analyses; analytic code; any other materials used in the review. | Data availability |

*From:* Page MJ, McKenzie JE, Bossuyt PM, Boutron I, Hoffmann TC, Mulrow CD, et al. The PRISMA 2020 statement: an updated guideline for reporting systematic reviews. BMJ 2021;372:n71. doi: 10.1136/bmj.n71. This work is licensed under CC BY 4.0. To view a copy of this license, visit https://creativecommons.org/licenses/by/4.0/

### Supplementary Table S2. Search strategies for each database

#### Supplementary Table S2a. PubMed

| Category | Search term |
|---|---|
| #1 (Disease) | "nipah"[All Fields] OR "nipa*"[All Fields] OR ("henipavirus"[MeSH Terms] OR "henipavirus"[All Fields] OR "henipaviruses"[All Fields]) OR "henipa"[All Fields] OR "henipa*"[All Fields] OR ("nipah virus"[MeSH Terms] OR ("nipah"[All Fields] AND "virus"[All Fields]) OR "nipah virus"[All Fields]) OR ("henipavirus infections"[MeSH Terms] OR ("henipavirus"[All Fields] AND "infections"[All Fields]) OR "henipavirus infections"[All Fields] OR ("nipah"[All Fields] AND "virus"[All Fields] AND "encephalitis"[All Fields]) OR "nipah virus encephalitis"[All Fields]) OR ("nipah"[All Fields] AND ("encephalities"[All Fields] OR "encephalitis"[MeSH Terms] OR "encephalitis"[All Fields])) OR ("niv"[All Fields] AND ("encephalities"[All Fields] OR "encephalitis"[MeSH Terms] OR "encephalitis"[All Fields])) |
| #2 (Epidemiology) | "epidemiologies"[All Fields] OR "epidemiology"[MeSH Subheading] OR "epidemiology"[All Fields] OR "epidemiology"[MeSH Terms] OR "epidemiology s"[All Fields] OR "epidemiolog*"[All Fields] OR ("disease outbreaks"[MeSH Terms] OR ("disease"[All Fields] AND "outbreaks"[All Fields]) OR "disease outbreaks"[All Fields] OR "outbreak"[All Fields] OR "epidemiology"[MeSH Subheading] OR "epidemiology"[All Fields] OR "outbreaks"[All Fields] OR "outbreak s"[All Fields]) OR "outbreak*"[All Fields] OR ("epidemiology"[MeSH Subheading] OR "epidemiology"[All Fields] OR "surveillance"[All Fields] OR "epidemiology"[MeSH Terms] OR "surveilance"[All Fields] OR "surveillances"[All Fields] OR "surveilled"[All Fields] OR "surveillence"[All Fields]) OR "cluster*"[All Fields] OR Reproduction number* OR R0 OR basic reproduction number* OR overdispersion |
| (#1 AND #2) | |

Number of results: 1,454

Date: Feb 28, 2026

#### Supplementary Table S2b. Embase

| Category | Search term |
|---|---|
| #1 (Disease) | exp Nipah virus/ or exp Nipah virus infection/ or nipah.mp. or nipah virus.mp. or nipah virus encephalitis.mp. or henipa*.mp. [mp=title, abstract, heading word, drug trade name, original title, device manufacturer, drug manufacturer, device trade name, keyword heading word, floating subheading word, candidate term word] |
| #2 (Epidemiology) | (exp epidemic/ or epidemic.mp.) or (exp article/ or article.mp.) or (exp epidemiology/ or epidemiology.mp.) or (exp basic reproduction number/ or basic reproduction number.mp. or (exp disease transmission/ or disease transmission.mp.)) or (exp dispersion/ or dispersion.mp.) or cluster*.mp. |
| #1 AND #2 | |

Number of results: 2,240

Date: Feb 28, 2026

#### Supplementary Table S2c. Web of Science

| Category | Search term |
|---|---|
| #1 (Disease) | https://www.webofscience.com/wos/woscc/summary/642e2035-cc00-4da6-8ce3-3024cad8dec5-0147dca1c4/relevance/1 ALL=(nipah OR nipa* OR henipavirus OR nipah virus infection* OR nipah virus encephalitis OR niv encephalitis) |
| #2 (Epidemiology) | https://www.webofscience.com/wos/woscc/summary/776d9fa3-9ac5-48e5-9079-04a1c579e882-0147dcc14b/relevance/1 ALL=(epidemiology* OR epidemic OR outbreak* OR surveillance* OR cluster* OR reproduction number* OR basic reproduction number* OR overdispersion) |
| #1 AND #2 | |

Number of results: 1,755

Date: Feb 28, 2026

### Supplementary Table S2d. Grey literature search

| WHO Archives of the Weekly Epidemiological Record | WHO Disease Outbreak News (Link) |
|---|---|
| Search term = ”nipah” | Search term = “nipah” |
| Number of results = 8 | Number of results = 13 |

Date: Feb 28, 2026

### Supplementary Table S3. Inclusion and exclusion criteria

#### Supplementary Table S3a. Title and abstract screening criteria

| **Include** if the title or abstract indicates at least one of the following keywords (or their synonyms) | **Exclude** if the title or abstract indicates that |
|---|---|
| Nipah virus | The study involves animals only OR |
| Nipah encephalitis | The study is a modelling study OR |
| Epidemiological parameter of Nipah virus | The study is a review or a systematic literature review |

#### Supplementary Table S3b. Full-text screening criteria

1) The same with title & abstract screening criteria
2) We will include studies with observational details of the outbreak; exclude studies without any outbreak-level information
3) We will record the details of the outbreak in the pre-defined data management spreadsheet
4) We will identify each outbreak by date and geography
5) If multiple studies describe the same outbreak, we prioritise
    a) The study that includes the most complete case ascertainment
    b) The study that provides the most complete details on the infector-infectee pairs

### Supplementary Table S4. List of data extracted

| Variable name | Type | Description |
|---|---|---|
| study_id | str | Study citation key. |
| doi | str | Digital Object Identifier. |
| outbreak_id | str | Outbreak/cluster identifier. |
| person_to_person | cat | 1 if involved person-to-person transmission at outbreak level; 0 if not. |
| pair_id | str | Pair ID for infector-infectee pair. |
| infector_id | str | Case ID for infector. |
| infectee_id | str | Case ID for infectee. |
| link_certainty | cat | link certainty based on the contact tracing result. |
| direction_notes | cat | notes on details of the transmission event. |
| setting | cat | setting of the transmission. |
| infectee_type | cat | isolated: spillover case which did not cause any secondary case; spillover_spread: spillover case which caused secondary case(s); sec_spread: secondary case which caused secondary case; sec_non_spread: secondary case which did not cause any further case; sec_spread_multiple_candidate: secondary case which caused secondary case(s) and one of the multiple infector candidates |
| exposure_event_type | cat | type of the exposure event based on the text description. |
| exposure_env | cat | environment of the exposure based on the text description. |
| contact_intensity | cat | contact intensity based on the text description. |
| ppe_use | cat | Personal Protection Equipment use based on the text description. |
| mask_use | str | Mask use based on the text description. |
| country_iso3 | str | ISO-3166-1 alpha-3 code. |
| admin1 | str | First-level admin area. |
| admin2 | str | Second-level admin area. |
| admin3 | str | Third-level admin area. |
| exposure_start_lower | date | Exposure window start (lower). |
| exposure_start_upper | date | Exposure window start (upper). |
| exposure_end_lower | date | Exposure window end (lower). |
| exposure_end_upper | date | Exposure window end (upper). |
| exposure_date_precision | str | notes on exposure date precision. |

| | | |
|---|---|---|
| exposure_duration | str | notes on exposure duration. |
| proximity_meters | str | notes on proximity distance. |
| infector_onset_lower | date | Infector symptom onset (lower). |
| infector_onset_upper | date | Infector symptom onset (upper). |
| infectee_onset_lower | date | Infectee symptom onset (lower). |
| infectee_onset_upper | date | Infectee symptom onset (upper). |
| infector_onset_precision | cat | Precision of the reported date of symptom onset of infector. |
| infectee_onset_precision | cat | Precision of the reported date of symptom onset of infectee. |
| multiple_candidate_infectors | int | Count of plausible infectors for infectee. |
| infector_outcome | cat | recorded outcome of the infectee; NA if infector is not a person. |
| infectee_outcome | cat | recorded outcome of the infectee. |
| infector_death_date | date | recorded date of death of the infector |
| infectee_death_date | date | recorded date of death of the infectee |
| notes | str | free notes |
| location_detail | str | notes on location further than admin level 3 |

### Supplementary Table S5. Studies included in the systematic review

| First author (Year) | Country | Outbreak year | Region | Cases (deaths) | Person-to-person transmission | Case-level data extraction |
|---|---|---|---|---|---|---|
| Chadha et al. (2006) (1) | India | 2001 | Siliguri (West Bengal) | 66 (45) | Yes | No* |
| Harit et al. (2006) (2) | India | 2001 | Siliguri (West Bengal) | 66 (45) | Yes | Yes* |
| Arankalle et al. (2011) (36) | India | 2007 | Nadia (West Bengal) | 5 (5) | Yes | Yes |
| Arunkumar et al. (2019) (14) | India | 2018 | Kozhikode (Kerala) | 23 (21) | Yes | Yes* |
| Pallivalappil et al. (2020) (37) | India | 2018 | Kozhikode (Kerala) | 23 (21) | Yes | No* |
| Ramachandran et al. (2022) (38) | India | 2019 | Ernakulam (Kerala) | 1 (0) | No | Yes |
| Yadav et al. (2022) (39) | India | 2021 | Kozhikode (Kerala) | 1 (1) | No | Yes |
| As et al. (2024) (40) | India | 2023 | Kozhikode (Kerala) | 6 (2) | Yes | Yes |
| Sahay et al. (2025) (41) | India | 2024 | Malappuram (Kerala) | 2 (2) | No | Yes |
| WHO DONs† (2025) (42) | India | 2025 | Malappuram (Kerala) | 2 (1) | No | Yes |
| | | 2025 | Palakkad (Kerala) | 2 (1) | No | Yes |
| WHO DONs† (2026) (19) | India | 2025 | North 24 Parganas (West Bengal) | 3 (2) | Yes | Yes* |
| WHO SEARO report (20) | India | 2025 | North 24 Parganas (West Bengal) | 3 (2) | Yes | Yes* |
| Hsu et al. (2004) (21) | Bangladesh | 2001 | Meherpur | 13 (9) | Yes | Yes* |
| | | 2003 | Naogaon | 12 (8) | Yes | Yes* |
| icddr,b HSB‡ (2003) (4) | Bangladesh | 2001 | Meherpur | 13 (9) | Yes | No* |
| | | 2003 | Naogaon | 12 (8) | Yes | No* |
| Montgomery et al. (2008) (3) | Bangladesh | 2004 | Rajbari | 12 (10) | No | Yes* |
| Gurley et al. (2007) (6) | Bangladesh | 2004 | Faridpur | 36 (27) | Yes | Yes* |
| icddr,b HSB‡ (2004) (4) | Bangladesh | 2004 | 8 districts§ | 29 (22) | No | No* |
| WHO WER¶ (2004) (5) | Bangladesh | 2004 | 6 districts** | 23 (17) | Yes | No* |
| | | 2004 | Faridpur | 30 (18) | Yes | No* |
| Luby et al. (2006) (7) | Bangladesh | 2004–2005 | Tangail | 12 (11) | No | Yes* |
| icddr,b HSB‡ (2005) (4) | Bangladesh | 2004–2005 | Tangail | 12 (11) | No | No* |
| Homaira et al. (2010)a (8) | Bangladesh | 2007 | Thakurgaon | 7 (3) | Yes | Yes* |
| Homaira et al. (2010)b (9) | Bangladesh | 2007 | Kushtia | 8 (5) | Yes | Yes* |
| icddr,b HSB‡ (2007) (4) | Bangladesh | 2007 | Thakurgaon | 7 (3) | Yes | No* |
| | | 2007 | Kushtia | 8 (5) | Yes | No* |
| Rahman et al. (2012) (43) | Bangladesh | 2008 | Manikganj | 4 (4) | No | Yes |
| | | 2008 | Rajbari | 6 (5) | No | Yes |

| | | | | | | |
|---|---|---|---|---|---|---|
| Sazzad et al. (2013) (22) | Bangladesh | 2010 | Faridpur | 16 (14) | Yes | Yes |
| icddr,b HSB‡ (2011) (4) | Bangladesh | 2011 | Lalmonirhat | 20 (20) | Yes | No* |
| icddr,b HSB‡ (2012) (4) | Bangladesh | 2011 | Rangpur | 7 (4) | Yes | Yes* |
| Chakraborty et al. (2016) (15) | Bangladesh | 2011 | Lalmonirhat | 22 (21) | Yes | Yes* |
| | | 2011 | Rangpur | 8 (5) | Yes | Yes* |
| | | 2011 | Dinajpur | 5 (4) | Yes | Yes |
| | | 2011 | Rajbari | 2 (2) | Yes | Yes |
| | | 2011 | Nipah-belt districts and Comilla†† | 6 (6) | No | Yes |
| Islam et al. (2016) (16) | Bangladesh | 2011 | Rangpur | 7 (4) | Yes | No* |
| | | 2012 | Rajshahi | 3 (2) | Yes | Yes |
| | | 2014 | Rangpur | 4 (2) | Yes | Yes |
| Hassan et al. (2018) (44) | Bangladesh | 2013–2014 | Faridpur, Rajshahi, Rangpur‡‡ | 16 (13) | Yes | Yes |
| Rahman et al. (2025) (17) | Bangladesh | 2023 | Rajbari | 3 (2) | Yes | Yes* |
| WHO DONs† (2023) (18) | Bangladesh | 2023 | Rajbari | 3 (2) | Yes | No* |
| | | 2023 | 7 districts§§ | 9 (7) | No | Yes |
| WHO DONs† (2024) (45) | Bangladesh | 2024 | Manikonj | 1 (1) | No | Yes |
| | | 2024 | Shariatpur | 1 (1) | No | Yes |
| WHO DONs† (2025) (46) | Bangladesh | 2025 | Pabna | 1 (1) | No | Yes |
| | | 2025 | Bhola | 1 (1) | No | Yes |
| | | 2025 | Faridpur | 1 (1) | No | Yes |
| | | 2025 | Naogaon | 1 (1) | No | Yes |
| WHO DONs† (2026) (47) | Bangladesh | 2026 | Naogaon | 1 (1) | No | Yes |

* Details on choosing studies describing the same outbreaks are provided in Supplementary Text S1.
† World Health Organization. Disease Outbreak News.
‡ icddr,b Health and Science Bulletin.
§ Rajbari, Joypurhat, Naogaon, Natore, Faridpur, Gopalganj, Manikganj, Dhaka
¶ World Health Organization. Weekly Epidemiological Record.
** Faridpur, Golpagonj, Maknikgonj, Joypurat, Naogaon, Rajbari.
†† First NiV case in the eastern area of Bangladesh.
‡‡ Consisted of four clustered outbreaks and one sporadic outbreak.
§§ Rajbari, Narsingdi, Shariatpur, Naogaon, Natore, Pabna, Rajshahi.

### Supplementary Table S6. Quality assessment results

#### Supplementary Table S6a. Case-control series

| Author | Year | Q1 | Q2 | Q3 | Q4 | Q5 | Q6 | Q7 | Q8 | Q9 | Q10 | Total Yes | Score (/10) | Quality |
|---|---|---|---|---|---|---|---|---|---|---|---|---|---|---|
| Hsu et al. | 2004 | Y | U | Y | U | Y | U | N | Y | Y | Y | 6 | 6/10 | Moderate |
| Luby et al. | 2006 | Y | Y | Y | Y | Y | U | U | Y | Y | Y | 8 | 8/10 | High |
| Gurley et al. | 2007 | Y | Y | Y | U | Y | Y | Y | Y | Y | Y | 9 | 9/10 | High |
| Montgomery et al. | 2008 | Y | U | Y | U | Y | U | U | U | Y | Y | 5 | 5/10 | Moderate |
| Homaira et al. | 2010a | Y | U | Y | U | Y | U | U | U | Y | Y | 5 | 5/10 | Moderate |
| Homaira et al. | 2010b | Y | U | Y | U | Y | U | U | U | Y | Y | 5 | 5/10 | Moderate |
| Rahman et al. | 2012 | Y | U | Y | Y | Y | U | U | U | Y | Y | 6 | 6/10 | Moderate |
| Sazzad et al. | 2013 | Y | U | Y | U | Y | U | U | U | Y | Y | 5 | 5/10 | Moderate |
| Chakraborty et al. | 2016 | Y | Y | Y | U | Y | Y | Y | U | Y | Y | 8 | 8/10 | High |

Y = Yes. N = No. U = Unclear. NA = Not Applicable. Q1: Were the groups comparable other than the presence/absence of disease? Q2: Were cases and controls matched appropriately? Q3: Were the same criteria used for identification of cases and controls? Q4: Was exposure measured in a standard, valid and reliable way? Q5: Was exposure measured in the same way for cases and controls? Q6: Were confounding factors identified? Q7: Were strategies to deal with confounding factors stated? Q8: Were outcomes assessed in a standard, valid and reliable way? Q9: Was the exposure period of interest long enough to be meaningful? Q10: Was appropriate statistical analysis used? Quality: Low < 5; 5 ≤ Moderate < 7; High ≥ 7

#### Supplementary Table S6b. Case series studies

| Author | Year | Q1 | Q2 | Q3 | Q4 | Q5 | Q6 | Q7 | Q8 | Q9 | Q10 | Total Yes | Score (/10) | Quality |
|---|---|---|---|---|---|---|---|---|---|---|---|---|---|---|

| | | | | | | | | | | | | | | |
|---|---|---|---|---|---|---|---|---|---|---|---|---|---|---|
| Chadha et al. | 2006 | Y | Y | Y | U | U | Y | Y | Y | Y | Y | 8 | 8/10 | High |
| Harit et al. | 2006 | Y | Y | Y | U | U | Y | Y | Y | Y | Y | 8 | 8/10 | High |
| Arankalle et al. | 2011 | U | Y | Y | U | U | U | U | U | Y | Y | 4 | 4/10 | Low |
| Islam et al. | 2016 | Y | Y | Y | U | Y | Y | Y | Y | Y | Y | 9 | 9/10 | High |
| Hassan et al. | 2018 | Y | Y | Y | U | U | Y | Y | Y | Y | Y | 8 | 8/10 | High |
| Arunkumar et al. | 2019 | Y | Y | Y | Y | Y | Y | Y | Y | Y | Y | 10 | 10/10 | High |
| Pallivalappil et al. | 2020 | Y | Y | Y | Y | Y | Y | Y | Y | Y | Y | 10 | 10/10 | High |
| Ramachandran et al. | 2022 | Y | Y | Y | U | U | Y | Y | Y | Y | U | 7 | 7/10 | High |
| As et al. | 2024 | Y | Y | Y | Y | Y | Y | Y | Y | Y | Y | 10 | 10/10 | High |
| Sahay et al. | 2025 | Y | Y | Y | U | U | Y | Y | Y | Y | Y | 8 | 8/10 | High |

Y = Yes. N = No. U = Unclear. NA = Not Applicable. Q1: Clear inclusion criteria? Q2: Condition measured in standard, reliable way? Q3: Valid methods for identification of condition? Q4: Consecutive inclusion of participants? Q5: Complete inclusion of participants? Q6: Clear reporting of participant demographics? Q7: Clear reporting of clinical information? Q8: Outcomes/follow-up results clearly reported? Q9: Clear reporting of presenting site/clinic demographics? Q10: Statistical analysis appropriate? Quality: Low < 5; 5 ≤ Moderate < 7; High ≥ 7

### Supplementary Table S6c. Case reports

| Author | Year | Q1 | Q2 | Q3 | Q4 | Q5 | Q6 | Q7 | Q8 | Total Yes | Score (/8) | Quality |
|---|---|---|---|---|---|---|---|---|---|---|---|---|
| Yadav et al. | 2022 | Y | Y | Y | Y | Y | Y | Y | Y | 8 | 8/8 | High |
| Rahman et al. | 2025 | Y | Y | Y | Y | Y | Y | Y | Y | 8 | 8/8 | High |

Y = Yes. N = No. U = Unclear. NA = Not Applicable. Q1: Patient demographic characteristics clearly described? Q2: Patient history described and presented as timeline? Q3: Current clinical condition on presentation clearly described? Q4: Diagnostic tests/methods and results clearly described? Q5: Intervention(s) or treatment procedure(s) clearly described? Q6: Post-intervention clinical condition clearly described? Q7: Adverse events (harms) or unanticipated events described? Q8: Case report provides takeaway lessons? Quality: Low < 5; 5 ≤ Moderate < 7; High ≥ 7

Results: *Overall.* Among case-control studies (n = 9), JBI critical appraisal scores (48) ranged 5 to 9 out of 10 (mean 6.3); three were rated high quality and six moderate (3,6–9,15,21,22,43). Case series (n = 10) scored 4–10 (mean 8.2); nine were high quality (1,2,14,16,36–38,40,41,44). The exception, Arankalle et al. (2011), scored 4/10 owing to its primary focus on genomic characterisation, with limited epidemiology-related information (36). Both case reports (n = 2) scored 8/8 (17,39).

*Case-control studies.* The nine case-control studies scored between 5 and 9 out of 10 (mean 6.3), with three rated High quality (Luby et al., 2006; Gurley et al., 2007; Chakraborty et al., 2016) and six Moderate (Hsu et al., 2004; Montgomery et al., 2008; Homaira et al., 2010a, 2010b; Rahman et al., 2012; Sazzad et al., 2013). All nine studies met criteria on Q1 (group comparability), Q3 (consistent case and control identification criteria), Q5 (exposure measured identically across groups), Q9 (adequate exposure period), and Q10 (appropriate statistical analysis). Four items showed widespread deficiencies. Appropriate matching (Q2) was confirmed in only three studies (Luby et al., 2006; Gurley et al., 2007; Chakraborty et al., 2016); the remaining six provided insufficient detail to confirm matching procedures. Validity and reliability of exposure measurement (Q4) was rated Yes in only two studies (Luby et al., 2006; Rahman et al., 2012), with the majority relying on retrospective recall interviews without formal validation. Confounding identification (Q6) and strategies to address confounding (Q7) were each met by two studies only (Gurely et al., 2007; Chakraborty et al., 2016); Hsu et al. (2004) was the sole study scored No on Q7. Outcome assessment validity (Q8) was confirmed in three studies (Hsu et al., 2004; Luby et al., 2006; Gurley et al., 2007), while six studies provided insufficient methodological detail to judge this item.

*Case series.* The ten case series scored between 4 and 10 out of 10 (mean 8.2), with nine rated High quality and one Low (Arankalle et al, 2011). Condition measurement (Q2), validity of identification methods (Q3), and reporting of presenting site characteristics (Q9) were met by all ten studies. Reporting of participant demographics (Q6), clinical information (Q7), and outcomes (Q8) was satisfactory in nine studies, with Arankalle et al. (2011) the exception across all three items, reflecting its primary focus on genomic characterisation rather than clinical or epidemiological reporting. The most common deficiencies were consecutive inclusion (Q4), confirmed in only three studies (Arunkumar et al. 2019; Pallivalappil et al., 2020; As et al., 2024), and complete inclusion (Q5), confirmed in four (Islam et al, 2016; Arunkumar et al., 2019; Pallivalappil et al., 2020; As et al., 2024). A discernible improvement in methodological quality is apparent over time: all studies published from 2019 onwards scored 7 or above, and three achieved the maximum score of 10/10, compared with scores of 4–8 among studies published before 2019. This trajectory likely reflects the adoption of pre-defined case definitions and more systematic outbreak investigation approach in recent years.

*Case reports.* Both case reports (Yadav et al., 2022; Rahman et al., 2025) scored 8/8 (High quality), meeting all JBI criteria including clear demographic and clinical description, diagnostic detail, treatment reporting, outcome description, and provision of takeaway lessons.

### Supplementary Table S7. Person-to-person outbreaks by transmission chain

| Chain depth | Generations | Bangladesh (N outbreaks) | India (N outbreaks) | Total (N outbreaks) |
|---|---|---|---|---|
| 1 | 2 | 14 | 3 | 17 |
| 2 | 3 | 1 | 2 | 3 |
| 4 | 5 | 1 | 0 | 1 |
| Total | – | 16 | 5 | 21 |

Chain depth, the maximum number of sequential person-to-person transmission events from the index case (Generation 0); Generations, number of generations spanned (e.g., Generation 0 and Generation 1 = two generations); N outbreaks, number of outbreaks. No outbreaks reached a chain depth of three.

### Supplementary Table S8. Prior sensitivity

| Prior set | *R* (median) [95% CrI] | *R* (mean) [95% CrI] | $\sigma_R$ [95% CrI] | *k* [95% CrI] |
|---|---|---|---|---|
| P1: Weakly informative | 0.52 [0.29–1.04] | 0.56 [0.31–1.24] | 0.27 [0.01–0.98] | 0.04 [0.02–0.06] |
| P2: Epidemiologically informed, permissive heterogeneity | 0.43 [0.26–0.75] | 0.47 [0.28–0.90] | 0.28 [0.01–1.00] | 0.04 [0.03–0.06] |
| P3: Epidemiologically informed, conservative heterogeneity | 0.44 [0.27–0.74] | 0.45 [0.28–0.76] | 0.15 [0.01–0.49] | 0.04 [0.03–0.06] |

*R* (median), median reproduction number across outbreaks, exp($\mu_R$); *R* (mean), mean reproduction number across outbreaks, exp($\mu_R + \sigma_R{}^2/2$); $\sigma_R$, standard deviation of log($R_j$) across outbreaks, quantifying between-outbreak heterogeneity; *k*, dispersion parameter, with smaller values indicating greater case-level transmission heterogeneity. CrI, credible interval (2.5th–97.5th posterior percentile).

### Supplementary Table S9. Country-specific analyses: P1–P3

| Prior | Country | Outbreaks | Cases | *R* (median) [95% CrI] | *R* (mean) [95% CrI] | $\sigma_R$ [95% CrI] | *k* [95% CrI] |
|---|---|---|---|---|---|---|---|
| P1 | Bangladesh | 54 | 211 | 0.42 [0.21–0.94] | 0.47 [0.24–1.31] | 0.38 [0.02–1.23] | 0.05 [0.03–0.09] |
| P1 | India | 13 | 112 | 0.84 [0.30–3.04] | 1.01 [0.35–6.50] | 0.43 [0.02–1.64] | 0.03 [0.01–0.06] |
| P2 | Bangladesh | 54 | 211 | 0.35 [0.19–0.63] | 0.40 [0.23–0.87] | 0.39 [0.02–1.24] | 0.05 [0.03–0.09] |
| P2 | India | 13 | 112 | 0.47 [0.22–0.98] | 0.55 [0.27–1.75] | 0.42 [0.02–1.59] | 0.03 [0.02–0.06] |
| P3 | Bangladesh | 54 | 211 | 0.36 [0.21–0.65] | 0.37 [0.22–0.68] | 0.17 [0.01–0.53] | 0.05 [0.03–0.08] |
| P3 | India | 13 | 112 | 0.48 [0.24–0.97] | 0.49 [0.25–1.00] | 0.16 [0.01–0.54] | 0.03 [0.02–0.06] |

Independent hierarchical negative binomial fits by country-specific data subset and prior set (P1–P3). *R* (median), median reproduction number across outbreaks, exp($\mu_R$); *R* (mean), mean reproduction number across outbreaks, exp($\mu_R + \sigma_R{}^2/2$); $\sigma_R$, standard deviation of log($R_j$) across outbreaks, quantifying between-outbreak heterogeneity;

$k$, dispersion parameter, with smaller values indicating greater case-level transmission heterogeneity. CrI, credible interval (2.5th–97.5th posterior percentile).

### Supplementary Table S10. Offspring distribution sensitivity analyses (SA-O-CV)

| Description | Outbreaks | Cases | $R$ (median) [95% CrI] | $\psi$ [95% CrI] | $k$ at $R_{med}$ [95% CrI] | $\sigma_R$ [95% CrI] |
|---|---|---|---|---|---|---|
| Shared $R/k$ ratio ($\psi$), all outbreaks | 67 | 323 | 0.40 [0.25–0.66] | 10.22 [5.90–19.08] | 0.04 [0.02–0.06] | 0.24 [0.01–0.72] |

SA-O-CV: sensitivity analysis in which the ratio $\psi$ = $R/k$ was held constant across outbreaks. $R$ (median), median reproduction number across outbreaks, $\exp(\mu_R)$; $\psi$, shared ratio $R/k$ across outbreaks; $k$ at $R_{med}$, implied dispersion parameter at the median $R$ (= $R_{med}/\psi$), shown for direct comparability with the primary model's shared $k$ (0.04 [95% CrI: 0.03–0.06]); $\sigma_R$, standard deviation of $\log(R_j)$ across outbreaks, quantifying between-outbreak heterogeneity; CrI, credible interval (2.5th–97.5th posterior percentile).

### Supplementary Table S11. Derived dispersion quantities from primary model

| Quantity | Median | 95% CrI | Source | Description |
|---|---|---|---|---|
| $\psi$ | 10.87 | [5.72–22.00] | primary fit (derived) | $\psi$ = $R_{med}/k$: implied $R/k$ ratio at the median $R$ from primary model |
| CV at $R_{med}$ | 5.25 | [4.34–6.47] | primary fit (derived) | CV = sqrt(1/$R_{med}$ + 1/$k$): coefficient of variation of the negative binomial offspring distribution at $R$ (median) |

$\psi$ and CV are derived from the primary-model posterior draws without re-fitting, computed for each posterior draw and then summarised. For a negative binomial distribution with mean $R$ and dispersion parameter $k$, Var = $R$ + $R^2/k$, so CV = sqrt(Var)/$R$ = sqrt(1/$R$ + 1/$k$), evaluated here at $R_j$ = $R_{med}$. CrI, credible interval (2.5th–97.5th posterior percentile).

### Supplementary Table S12. Serial interval: primary and sensitivity analyses

| Analysis | Pairs | Best distribution | Mean [95% CI], days | SD [95% CI], days | Empirical median | Empirical SD | ΔAIC gamma | ΔAIC lnorm | ΔAIC Weibull |
|---|---|---|---|---|---|---|---|---|---|
| Primary | 137 | gamma | 13.3 [12.8–13.8] | 3.3 [2.7–3.9] | 13.0 | 3.2 | **0.0** | 10.0 | 5.1 |
| SA-S1: Unambiguous pairs only | 134 | gamma | 13.4 [12.9–14.0] | 3.2 [2.6–3.8] | 13.0 | 3.1 | **0.0** | 9.9 | 7.2 |
| SA-S2a: Bangladesh pre-2007 | 48 | gamma | 12.8 [11.8–13.9] | 3.6 [2.4–4.6] | 13.0 | 3.6 | **0.0** | 3.1 | 5.5 |
| SA-S2b: Bangladesh ≥2007 | 22 | weibull | 12.6 [11.4–13.9] | 3.0 [1.9–3.7] | 13.0 | 3.1 | 4.1 | 6.6 | **0.0** |

SA-S1 excludes infector-infectee pairs with unresolved multiple candidate infectors. SA-S2a/b compares periods before (pre-2007) and after (≥2007) the implementation of national surveillance in Bangladesh. CI, 95% bootstrap percentile interval (fixed best-fitting distribution, 1,000 resamples); ΔAIC, Akaike Information Criterion difference versus best-fitting distribution (values ≤2 indicate comparable fit); Bold, Best distribution selected by minimum AIC on each subset's full dataset. The best-fitting family differed for the post-2007 subset (Weibull), but the gamma and Weibull means for that subset differ by less than 0.1 days, well inside the bootstrap interval.

### Supplementary Figure S1. Epidemic curves of Nipah virus outbreaks in Bangladesh and India

Each panel shows the daily number of cases by symptom-onset date for each outbreak included in the study, labelled by year and location. Case counts include confirmed, probable, and suspected cases with an available onset date. Single-case spillover outbreaks and outbreaks without recorded onset dates were excluded. Axis scales differ across panels. Onset dates were extracted from the source investigations listed in Table S5.

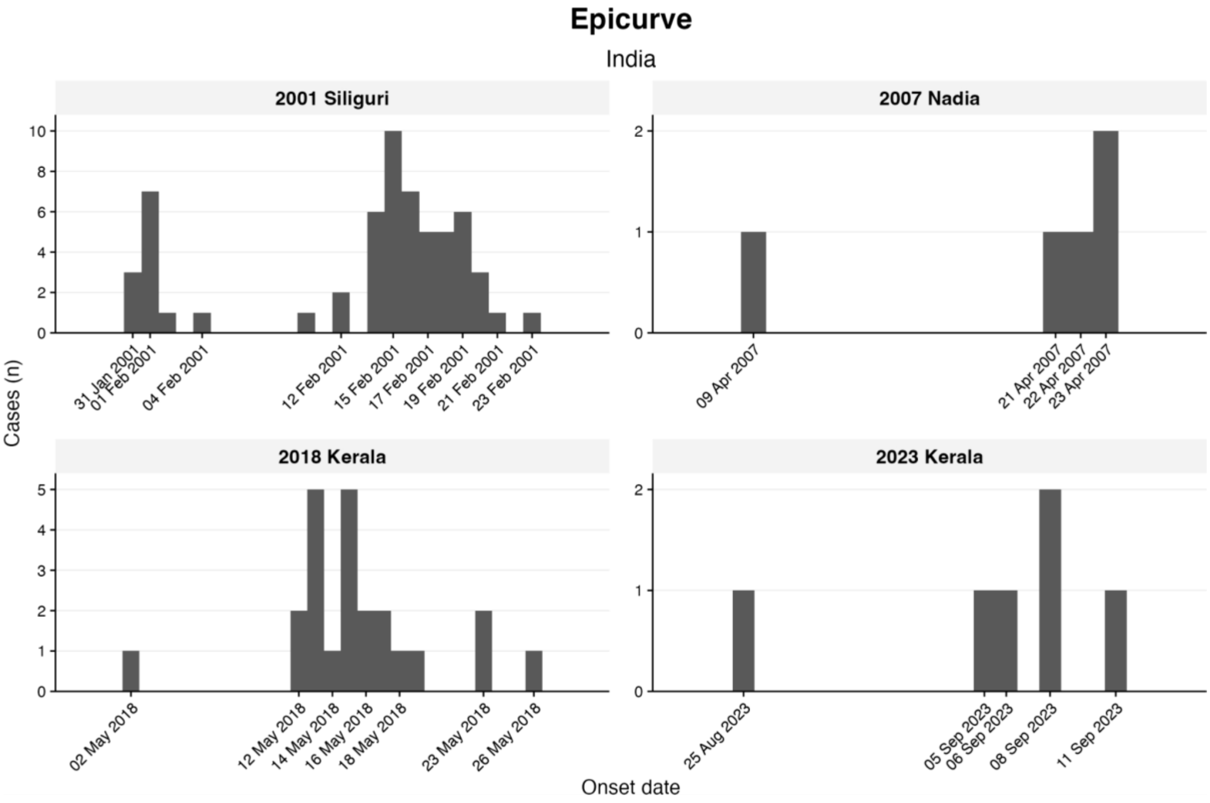

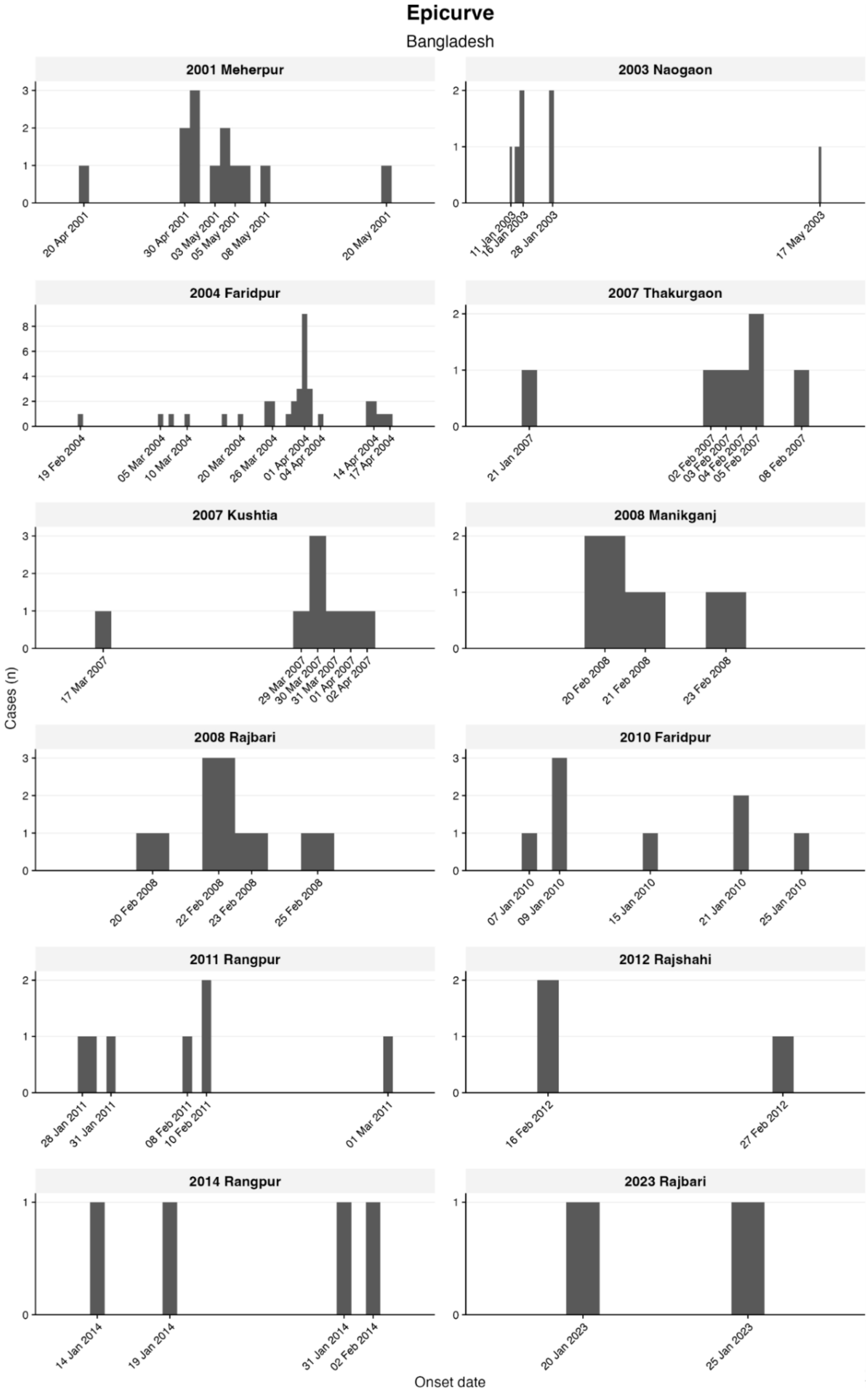
Epicurve
Bangladesh
2001 Meherpur
20 Apr 2001
30 Apr 2001
03 May 2001
05 May 2001
08 May 2001
20 May 2001
2003 Naogaon
11 Jan 2003
16 Jan 2003
28 Jan 2003
17 May 2003
2004 Faridpur
19 Feb 2004
05 Mar 2004
10 Mar 2004
20 Mar 2004
26 Mar 2004
01 Apr 2004
04 Apr 2004
14 Apr 2004
17 Apr 2004
2007 Thakurgaon
21 Jan 2007
02 Feb 2007
03 Feb 2007
04 Feb 2007
05 Feb 2007
08 Feb 2007
2007 Kushtia
17 Mar 2007
29 Mar 2007
30 Mar 2007
31 Mar 2007
01 Apr 2007
02 Apr 2007
2008 Manikganj
20 Feb 2008
21 Feb 2008
23 Feb 2008
2008 Rajbari
20 Feb 2008
22 Feb 2008
23 Feb 2008
25 Feb 2008
2010 Faridpur
07 Jan 2010
09 Jan 2010
15 Jan 2010
21 Jan 2010
25 Jan 2010
2011 Rangpur
28 Jan 2011
31 Jan 2011
08 Feb 2011
10 Feb 2011
01 Mar 2011
2012 Rajshahi
16 Feb 2012
27 Feb 2012
2014 Rangpur
14 Jan 2014
19 Jan 2014
31 Jan 2014
02 Feb 2014
2023 Rajbari
20 Jan 2023
25 Jan 2023
Cases (n)
Onset date

### Supplementary Figure S2. Prior and posterior densities under three prior specifications

Prior and posterior density of the median reproduction number ($R_{med}$), between-outbreak heterogeneity parameter ($\sigma_R$), and offspring dispersion parameter ($k$) under three prior specifications. P1: weakly informative. P2: epidemiologically informed, permissive heterogeneity. P3: epidemiologically informed, conservative heterogeneity. A: Pooled analysis (67 outbreaks, 323 cases). B: Bangladesh analysis (54 outbreaks, 211 cases). C: India analysis (13 outbreaks, 112 cases). Solid lines with shading: posterior densities. Dashed lines: prior densities, shown in grey where two specifications overlap (P2 and P3 for $R_{med}$ and $k$; P1 and P2 for $\sigma_R$). $\sigma_R$ is shown on a $\log_{10}$ axis, with densities with respect to $\log_{10}(\sigma_R)$. Vertical dotted line: posterior median under P2. Axes are truncated to the region of posterior support.

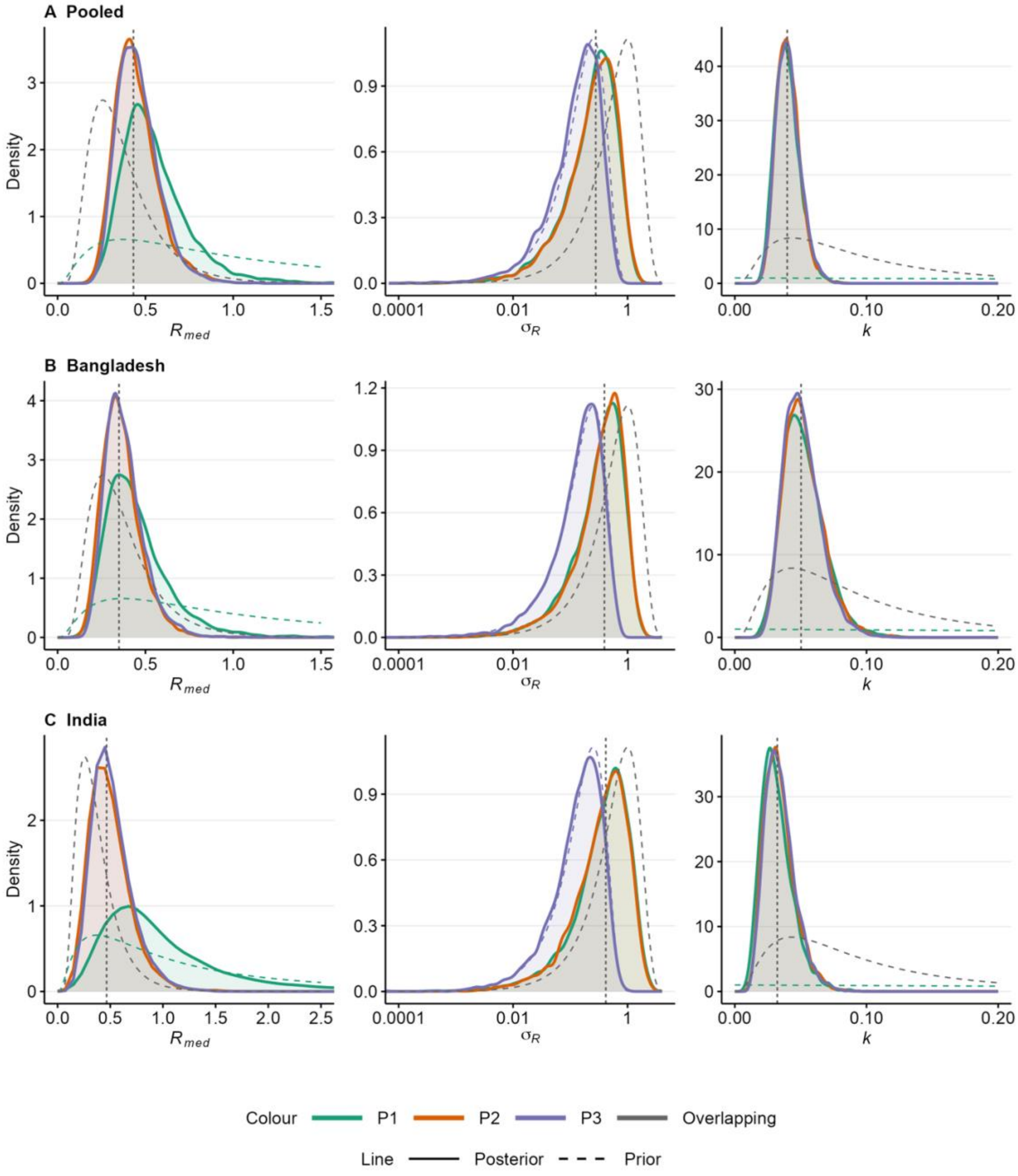

### Supplementary Figure S3. Posterior predictive check

Posterior predictive check for the primary model (P2, pooled analysis, n = 323 cases, 67 outbreaks). Histograms show distributions of 1,000 simulated values for each statistic. The red vertical line marks the observed value; dotted lines mark the 2.5th and 97.5th percentiles of the simulated distribution. Monte Carlo (MC) p-values are two-sided: values near 1 indicate the observed statistic is well-centred within the predictive distribution; values near 0 indicate poor fit. All three observed statistics fall within the 95% predictive intervals.

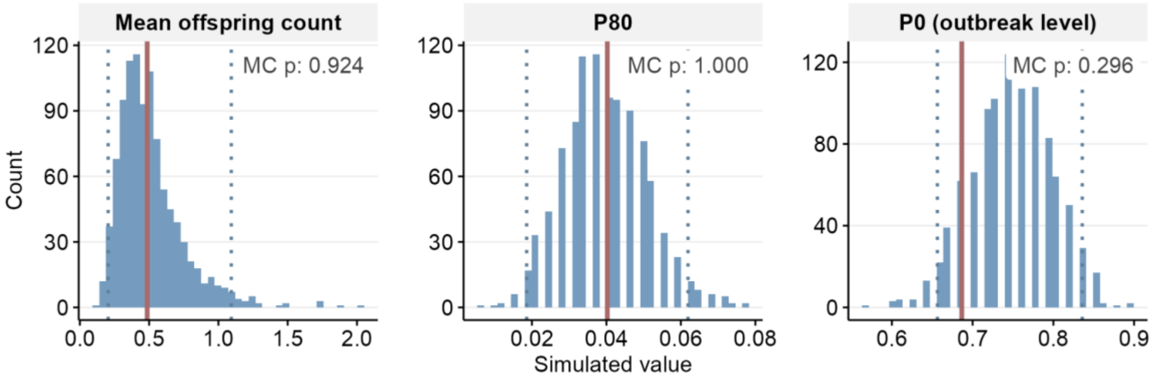

### Supplementary Figure S4. Observed and simulated offspring count distributions

Observed offspring count distribution (blue) versus marginal posterior predictive distribution (red). Red bars show the expected proportion at each secondary case count, averaged over 1,000 posterior predictive replicates from the primary model (P2). Values at x ≥ 15 are grouped. The marginal predictive distribution integrates over posterior uncertainty in outbreak-specific $R_j$ and $k$.

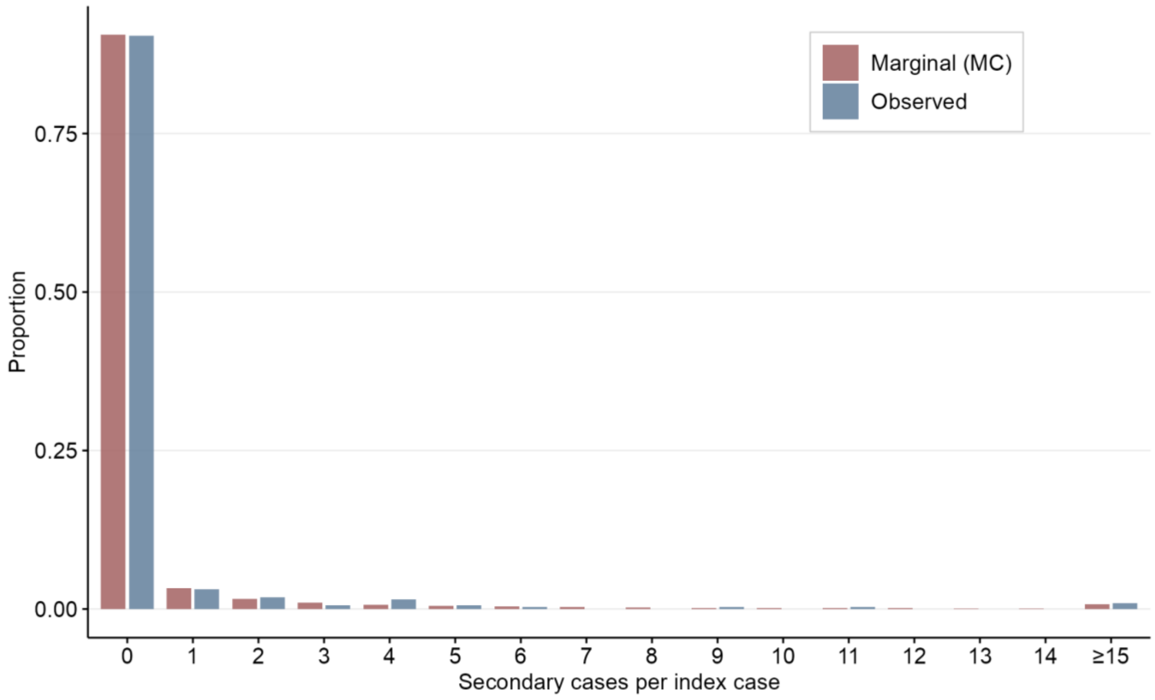

### Supplementary Figure S5. Trace plots

MCMC trace plots for population-level parameters from the primary P2 model. Four chains (different colours) show mixing without visible trends or between-chain separation. All diagnostic criteria met: $\hat{R} < 1.01$, effective sample size > 2,500, zero divergences.

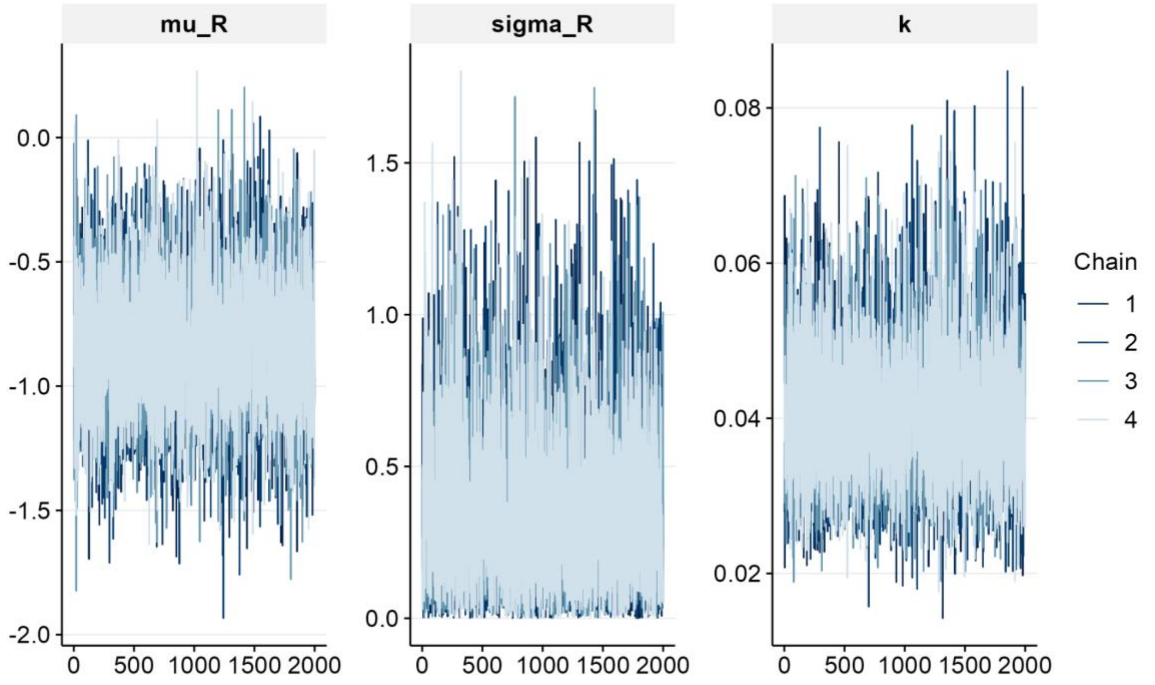

### Supplementary Figure S6. Posterior predictive check by country

Posterior predictive checks for country-specific models under P2. Top row: Bangladesh (54 outbreaks, 211 cases). Bottom row: India (13 outbreaks, 112 cases). Histograms show distributions of 1,000 simulated values for each statistic. The red vertical line marks the observed value; dotted lines mark the 2.5th and 97.5th percentiles of the simulated distribution. Monte Carlo (MC) p-values are two-sided: values near 1 indicate the observed statistic is well-centred within the predictive distribution; values near 0 indicate poor fit. All observed statistics fall within the 95% predictive intervals in both countries.

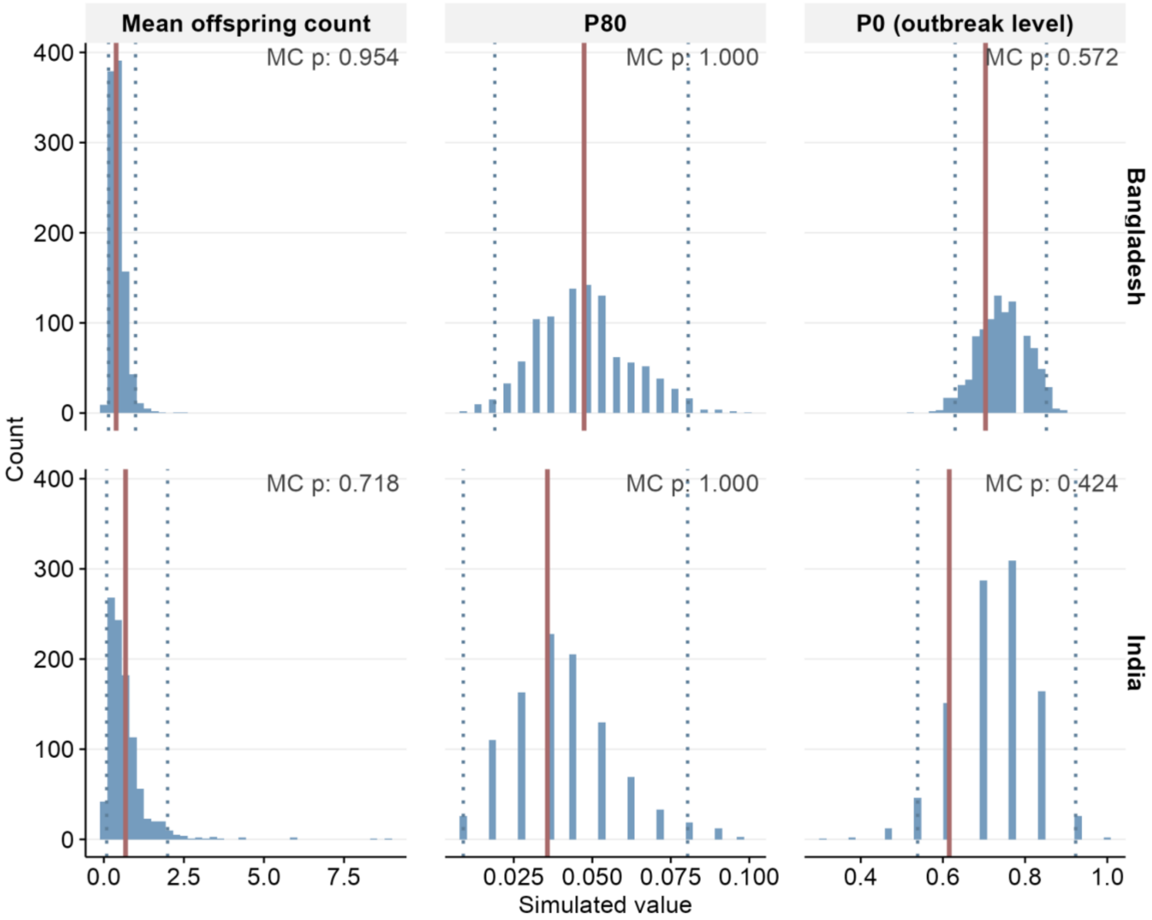

33. Naidoo T, Morgenstern C, Doohan P, Earl R, Rawson T, Sheppard RJ, et al. A systematic review of Nipah virus disease epidemiological parameters, outbreaks,

and mathematical models. Lancet Infect Dis. 2026 Oct;26(10):e409–22. doi:10.1016/S1473-3099(26)00239-2
